# Task-sharing echocardiographic screening for rheumatic heart disease in remote First Nations Australian communities: implementation evaluation from the NEARER SCAN study

**DOI:** 10.64898/2026.07.18.26358403

**Authors:** Ben Jones, Alice Mitchell, James Marangou, Jennifer Yan, Jeffrey Cannon, Jacqueline M Williamson, Lucy Law, Alex Kaethner, Meghan Bailey, Raelene Collins, Lucy Mayo, Vicki Wade, Dana Fitzsimmons, Amy Paterson, Bo Reményi, Anna P Ralph, Gavin Wheaton, Emma Haynes, Judith M Katzenellenbogen, Natasha J Howard, Peter Riley, Kurt Brown, Jessica Gatti, Sharon Lockyer, Chantelle Pears, Maida Stewart, Bronwyn Rossingh, Cassandra Daniels, Anferida Monteiro Fernades, Hilary Hardefeldt, Jessica O’Brien, Graham S Hillis, Daniel Engelman, Alex Brown, Andrew C Steer, Jonathan Carapetis, Mike English, Shobhana Nagraj, Joshua R Francis

## Abstract

**Background:** Rheumatic heart disease (RHD) remains a major cause of premature death in low- and middle-income countries and First Nations communities. Early detection and management can prevent progression, but requires echocardiography, which is limited in high-burden settings. Task-sharing echocardiographic screening is an accessible, evidence-based approach but implementation remains unclear.

**Methods:** We conducted a prospective implementation evaluation of a co-designed task-sharing screening programme across five remote First Nations Australian communities between May 2023 and November 2025. Predominantly community health workers (CHWs), alongside nurses and doctors, were trained to scan using handheld devices with off-site cardiologist interpretation. We assessed implementation outcomes and used a realist evaluation to explore how context shaped CHWs’ ability to complete training and embed screening into routine work. Data included scanning activity, surveys, costing, interviews, focus groups, and field notes.

**Findings:** We trained 32 staff (21 CHWs, 8 nurses, 3 doctors) to scan across five sites with 14 achieving certification. Scanning frequency was lower and more variable than anticipated: 360 scans (including training and post-certification) of 5–20-year-olds over 14 months, with site-level coverage of 3–85%. Fidelity was limited by device unavailability, charging problems, and delays in uploads and reviews. Set-up and training cost A$51,903/site, plus A$9,858/year in implementation support. Screening was easier for CHWs to embed when the legitimacy of their role as a scanner was communicated, but harder when invisible work outweighed opportunities to scan.

**Interpretation:** Future implementation will require efforts to legitimise CHWs’ scanning and support invisible work. Event-based screening offers a promising complementary strategy. Scale-up requires policy support.

**Funding:** This research was funded by the Australian Medical Research Futures Fund Cardiovascular Health Mission (GNT2015869), in addition to philanthropic donations from Medtronic Australasia, Edwards Life Sciences and the Rotary Club of Kiama. Hand-held devices (Philips Lumify, USA) were donated by Humpty Dumpty Foundation and East Timor Hearts Fund. BJ was supported by a Rhodes Scholarship.

**Research in context:** *Evidence before this study:* RHD is concentrated in low-resource settings where access to echocardiography is most limited. Previous studies have shown that task-sharing echocardiographic screening with briefly trained local scanners using handheld devices and simplified protocols to scan with off-site expert review, is an adequately accurate approach that could support early detection. However, the evidence has focused on diagnostic performance rather than implementation in routine health services. Little is known about implementation outcomes, cost, and the conditions required for screening to become embedded in everyday care.

*Added value of this study:* Our study has identified some of the real-world challenges in implementing this evidence-based early detection programme in practice, addressing a recognised gap for this prevalent disease of inequity. It advances understanding of how to implement task-sharing echocardiographic screening for RHD by identifying key implementation strategies, the mechanisms through which they operate, and the conditions that enable or hinder them. It quantifies what implementation costs in practice and identifies fidelity shortfalls. It extends the task-sharing literature by highlighting the importance of role legitimacy and invisible work when embedding a new practice into the routine care provided by CHWs.

*Implications of all the available evidence:* Task-sharing echocardiographic screening remains a community-supported and promising approach to early detection in remote First Nations Australian communities, but effective implementation requires more than initial training and devices. Ongoing sonographer visits, screening events, and implementation strategies that signal legitimacy and support invisible work will be needed to create the conditions for CHWs to scan regularly enough to maintain their skills and complete training in a timely manner. Policy support will be required, alongside further evaluation of long-term sustainability.

## 1 Background

Rheumatic heart disease (RHD) is the chronic damage to the heart valves caused by an abnormal immune response to group A streptococcal infections in childhood[1]. RHD affects an estimated 55 million people globally, resulting in 375,000 deaths each year[2]. It is a disease of inequity, with most cases occurring in low- and middle-income countries and First Nations communities, despite being largely eliminated in high-income settings by the 1980s[3].

Early detection with timely management can prevent disease progression[4]. However, reliable detection depends on skilled heart ultrasound (echocardiography), which can be difficult to access in high-burden settings. Echocardiographic screening conducted by trained local scanners using simplified imaging protocols on handheld devices offers an accessible approach, and is supported by accumulating evidence over the past decade[5, 6]. This approach is now included in the latest World Heart Federation guidelines for the echocardiographic diagnosis of RHD, reflecting growing expert consensus[7].

At the same time, the guidelines also highlight that strategies for implementation of echocardiographic screening are uncertain. It remains unclear whether screening can be implemented as intended in real-world practice, what coverage is achievable, what the cost drivers are, and which conditions support or hinder the embedding of screening into routine practice.

Addressing these gaps is timely in Australia, where a pilot health system–led screening programme, the National Aboriginal Community Controlled Health Organisation’s ‘Echo in ACCHOs’ programme, has commenced training in First Nations primary healthcare facilities, which pose challenging implementation environments[8, 9]. In Australia, the burden of RHD falls disproportionately on First Nations peoples, particularly those living in remote communities[10]. Some of these communities experience among the highest documented rates globally, with a prevalence in school-age children reaching 8% in one community[11]. Current models of detection largely rely on visiting specialist outreach services or hospitalisation at a tertiary centre following onset of symptoms. As a result, opportunities for early detection are frequently missed[12].

The ‘Non-Expert Acquisition and Remote Expert Review of Screening echocardiography images from Child health and AnteNatal clinics’ (NEARER SCAN) study involved the implementation of a task-sharing echocardiographic screening programme for RHD in partnership with First Nations communities, using a co-designed implementation plan[13]. Local First Nations community health workers (CHWs)1, nurses, and doctors, were trained to perform a simplified screening protocol using handheld echocardiography to scan 5-20 year-olds and pregnant women. Images were subsequently uploaded for off-site cardiologist interpretation.

The aim of this evaluation was to improve understanding of how to implement task-sharing echocardiographic screening for RHD in remote First Nations communities. Specifically, we sought to (1) describe the extent, variability, and resource requirements of implementation across five sites, and (2) explain how, why, for whom, and under what conditions screening was (or was not) embedded into CHWs’ routine practice. This evaluation focuses mainly on early implementation, particularly the challenges of training and establishing a workforce capable of delivering screening, reflecting the stage reached across sites during the evaluation period. Analyses of the programme’s clinical effectiveness, including data from sites in Timor-Leste and a regional Australian hospital, are ongoing and will be reported separately.

## 2 Methods

This manuscript is reported in accordance with the Standards for Reporting Implementation Studies (StaRI) statement[14] and the RAMESES II standards for realist evaluations[15] (Supplementary Materials 1 and 2).

### 2.1 Study design

To complement the NEARER SCAN hybrid trial’s planned effectiveness evaluation (see protocol[16]), we evaluated the process and success of programme delivery using implementation outcome metrics. A realist evaluation was also used to explain the context and underlying mechanisms that shaped the embedding of screening into CHWs’ routine practice. This prospective evaluation was conducted between May 2023 and November 2025.

Before implementation, a co-design phase[13] helped define the programme’s form in each delivery site (described below). The initial programme theory was also developed during this phase (Supplementary Material 3). The evaluation was conducted in three phases: Phase 1 (May 2023–October 2024), Phase 2 (November 2024–June 2025), Phase 3 (July–November 2025) (Figure 1).

**Fig. 1.**
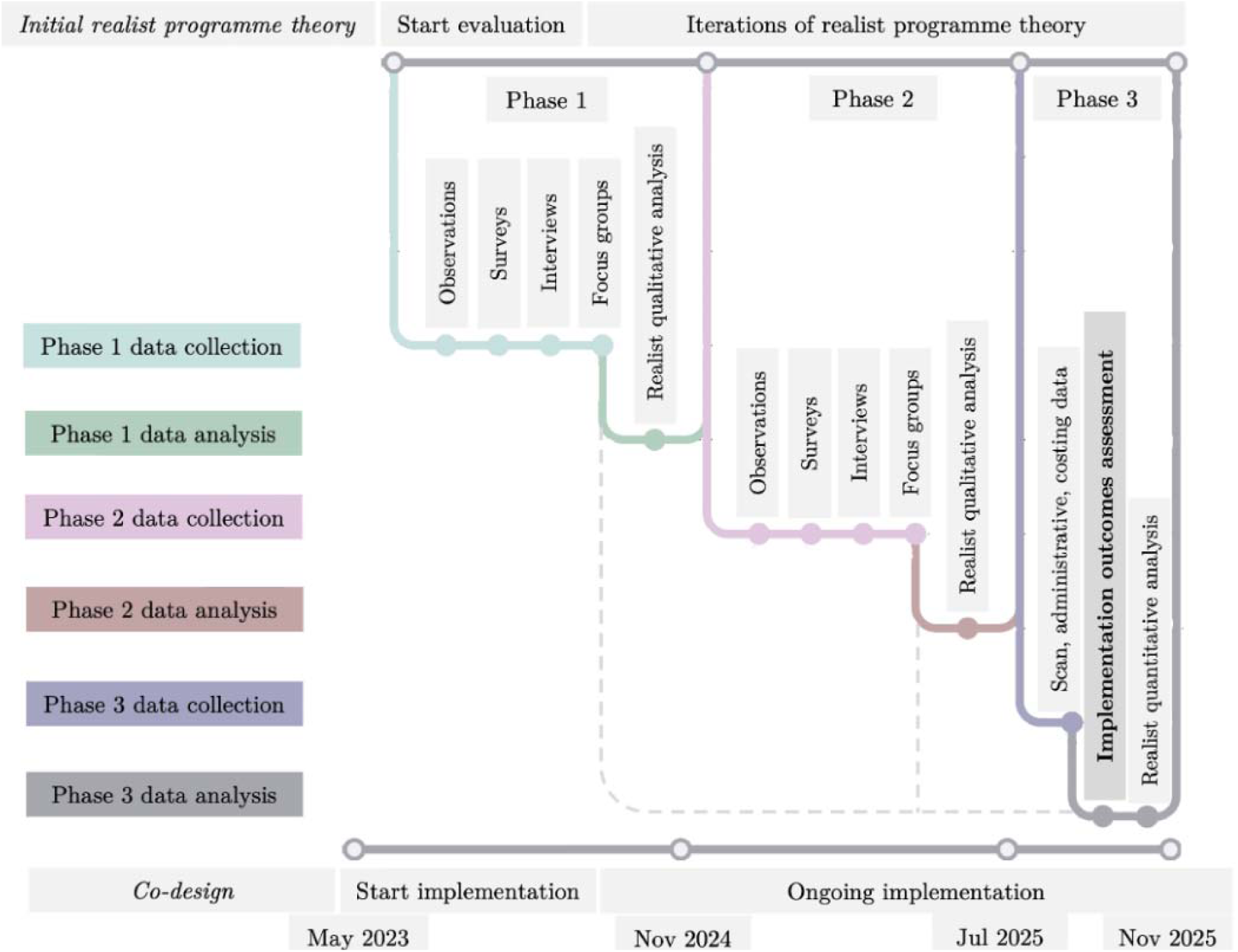
Evaluation design and timeline. Co-design and the development of the initial programme theory occurred prior to Phase 1 and are shown for context only. The initial programme theory informed the evaluation and was iteratively refined across three phases. Implementation outcomes were assessed in Phase 3, informed by data collected in earlier phases (dotted line indicates Phase 1–2 data informing this implementation outcomes assessment in Phase 3).

### 2.2 Study setting

The evaluation was conducted in five remote First Nations communities in Australia: one in the West Pilbara region of Western Australia and four in the Top End of the Northern Territory. Participating facilities were primary healthcare services, either government-run or Aboriginal Community Controlled Health Organisations (full details reported elsewhere[13, 16]). Given the small staff sizes at participating facilities, site names are represented numerically (Sites 1–5) to protect participant identity.

The primary cadre trained in handheld echocardiography were local First Nations CHWs, including qualified Aboriginal Health Practitioners, who require registration with the Australian Health Practitioner Regulation Agency, and community workers without formal qualifications employed by the local health facility. For all the CHWs trained, echocardiography represented a departure from usual scope of practice, although the extent of this varied. Some, particularly Aboriginal Health Practitioners, were already working in clinical roles, while others held more community-based roles and were being supported to take on expanded clinical responsibilities. Collectively, First Nations CHWs occupy a unique, often undervalued position within the Australian health workforce[17]. They bring community knowledge and language skills that improve facilities’ ability to provide culturally safe care. Although their knowledge and experience are well recognised within their communities, their formal authority in health services is often limited. For example, in one jurisdiction, some CHWs were only recently reclassified out of the same employment stream as cleaning staff, reflecting entrenched structural undervaluation[17]. They often experience role ambiguity, with considerable variation in scopes of practice across Australia[18]. In many facilities, CHWs serve as cultural brokers, and may be the only staff who speak local languages. These responsibilities shape their day-to-day work, as other staff rely on them for ad-hoc tasks such as patient transport. Our realist evaluation focused on CHWs because this aligns with community priorities for locally delivered, culturally safe, language-concordant care, and because they are the main cadre envisioned to deliver screening within a scaled-up programme[19].

### 2.3 Description of intervention

The screening programme comprised seven components aligned to a core programme function (Table 1) [20]. Further details are reported in Supplementary Material 4 using the Template for Intervention Description and Replication (TIDieR). Figure 2 provides an overview of the standardised screening programme and the site-specific implementation strategies used to support delivery across sites.

**Fig. 2.**
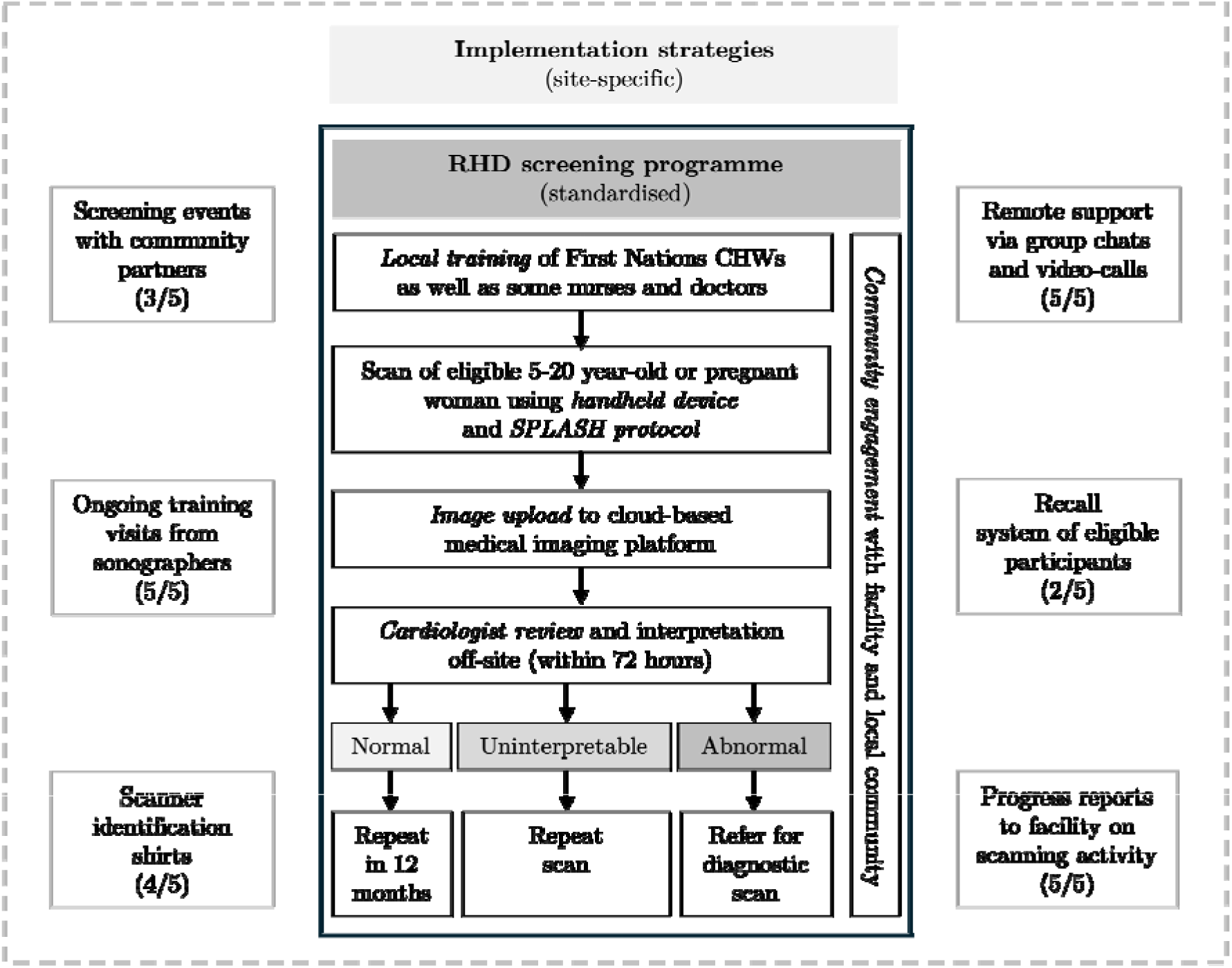
The RHD screening programme, standardised across sites, and example site-specific implementation strategies. /5 refers to the number of sites that implemented each example strategy.

**Table 1.** Programme components and functions.

| Component | Description | Core function |
| --- | --- | --- |
| Community engagement | Co-design a tailored implementation plan and iteratively adapt the programme in response to feedback[13]. | Maintain meaningful engagement between the programme and the broader community. |
| Local training | Four-days in-person (two theory, two practical) on Single Parasternal Long Axis view with a Sweep of the Heart (SPLASH) simplified echocardiographic protocol [21]. Delivered by experts for CHWs, nurses, and GPs, followed by 100 logbook scans and practical assessment. | Develop a trained local workforce capable of performing scans. |
| Handheld devices | Provision of tablets & Philips Lumify handheld probes (approximately one per two scanners). | Ensure functioning handheld devices are accessible for scans. |
| SPLASH protocol | Use of the evidence-based SPLASH protocol, aligned with World Heart Federation guidelines, which takes ≈ 10-minutes and can be performed through-the-collar (does not require removal of clothing) [22, 23]. | Acquire interpretable images using the SPLASH protocol. |
| Image upload | Transfer of studies from tablets to Tricefy, a secure cloud-based image platform, via the app on tablets provided. | Maintain a consistent and reliable image upload system to enable timely off-site review. |
| Cardiologist interpretation | Cardiologist interpret scans via Tricefy remotely within 72 hours, classify as normal, abnormal, or uninterpretable, provide feedback, and recommend a diagnostic echocardiogram (within 3 months), repeat screening, or management<sup>1</sup>. | Support timely off-site cardiologist interpretation and provide ongoing educational feedback to trained local staff on their scanning. |
| Care linkage<sup>2</sup> | Establish pathways linking abnormal screening results to routine clinical care, including referral for confirmatory echocardiography, and subsequent management. | Support timely follow-up and integration with routine clinical care. |

<sup>1</sup>Participants with markedly abnormal scans were occasionally initiated on treatment before confirmatory echocardiography.
<sup>2</sup>This programme component was not evaluated in this publication because, at this point of analysis, available data and follow-up time were insufficient to assess referral and downstream clinical care processes. A subsequent publication is planned to describe this component.

#### The NEARER SCAN training pathway

Trainees were nominated by facility managers, with preference given to local First Nations staff or to staff considered likely to remain at the facility longer term. Training comprised three stages. Stage I involved 4 days of in-person training delivered by expert sonographers or cardiologists, including 2 days of theory and 2 days of supervised scanning, with instruction focused on the Single Parasternal Long Axis view with a Sweep of the Heart (SPLASH) echocardiographic protocol[21]. Stage II required completion of 100 logbook scans, which could be performed on any consenting individual, regardless of screening eligibility. This stage was self-paced, with no fixed completion period. Certification was awarded following 100 logbook scans and a satisfactory in-person practical assessment. Stage III is an ongoing post-certification phase in which certified scanners screen only the eligible population and continue to receive supervised practice during periodic expert booster visits intended to reinforce skills.

### 2.4 Description of implementation strategies

Taking into account local context, an implementation plan was co-designed at each site prior to implementation[13]. Included implementation strategies were specified using the Expert Recommendations for Implementing Change (ERIC) taxonomy (Table 2)[24]. Responsibility for enacting strategies was shared between the implementation team and local staff, with the former leading training and supervision, and the latter leading workflow integration. The fidelity with which sites adopted the tailored package of strategies to achieve shared core functions was evaluated.

**Table 2.** Implementation strategies specified using the ERIC taxonomy.

| Implementation strategy | Description |
| --- | --- |
| Alter incentive structures | Celebrate completion of training with locally designed shirt identifying staff as a scanner. |
| Audit and provide feedback | Monthly scanning activity reports via email. |
| Capture and share local knowledge | Community-of-practice workshop (in-person, once-off) where scanners from different sites share experience. |
| Centralise technical assistance | Remote support via WhatsApp group with a designated sonographer for each site. |
| Change equipment | Provide internet dongles to address connectivity barriers for image upload. |
| Change service sites | Screening events delivered at local schools and community organisations. |
| Conduct ongoing training | Ongoing training visits to each site for in-person support. |
| Distribute educational materials | RHD brochures and posters promoting screening. |
| Identify and prepare champions | Lead to champion screening, assist with community engagement, coordinate activities, and troubleshoot barriers. |
| Make training dynamic | Adapt components of training to local language and preferences for experiential learning and storytelling. |
| Organise clinician implementation team meetings | Off-site support via regular video-calls. |
| Remind clinicians | Recall systems to prompt screening of eligible participants. |
| Shadow other experts | Pair new scanners with more experienced colleagues in a 'buddy' system, especially early after initial training. |
| Visit other sites | Facilitate opportunities to join regional mass screening week with expert cardiologists and sonographers. |

### 2.5 Data collection

Quantitative and qualitative data were collected to inform the evaluation (Tables 3 and 4). Relevant data were used to assess implementation outcomes (based on Proctor et al.’s taxonomy[25]) and support the evaluation where appropriate.

**Table 3.** Sources of data for implementation outcomes.

| Implementation outcome component [25] | Application in study | Data source |
| --- | --- | --- |
| Fidelity | *Intervention fidelity*: degree to which each programme component achieves its core function. *Implementation strategies* were delivered to support the achievement of core functions. Some were planned, while others were introduced and adapted throughout the study. | Theory of Change indicators, interviews, focus groups, observations, surveys, scan records, administrative data. |
| Adoption | Extent to which trainees commenced independent scanning in Stage II. Metrics: proportion of trainees who performed $\geq 1$ scans within 3 months of initial training (Stage I); median number of scans within the same 3 months; median time from Stage I training to first scan. | Scan records, administrative data. |
| Penetration | Extent to which scanning was delivered to the intended population. Metrics: total number of scans of 5–20 year-olds; percentage of eligible 5–20 year-olds scanned; proportion of weeks in which $\geq 1$ of eligible 5–20 year-olds had a scan. | Scan records, Australian Bureau of Statistics data. |
| Sustainability | Continued delivery of scanning throughout the study period. Metrics: proportion of weeks in which $\geq 1$ scan was uploaded during the final 3 months and facility staff's perceptions of the sustainability of the programme. | Scan records, administrative data. |

**Table 4.** Sources of data for realist evaluation.

| Realist evaluation component | Application in study | Data source |
| --- | --- | --- |
| Context | Specific conditions (e.g. individuals, interpersonal relations, institutions, and settings) that, when present, enable or constrain mechanisms to be triggered. | Interviews, focus groups, observations. |
| Mechanism | A hidden, context-sensitive, causal force. | Interviews, focus groups, observations. |
| Outcome | The intended and unintended change that occurs when a mechanism is triggered in a particular context. | Interviews, focus groups, observations, surveys, scan records, administrative data. |

#### 2.5.1 Primary data

Primary data collected during Phases 1 and 2 included interviews, focus groups, surveys, observations. Inclusion criteria for participating in these elements were being a staff member at a participating facility or a cardiologist reviewer. Participants were recruited using purposive sampling. Earlier sampling sought maximum variation, whereas later sampling targeted those best placed to refine emerging realist findings[26].

Semi-structured realist interviews were conducted by BJ and AM (example interview guide in Supplementary Material 5)[27]. Interviews were audio-recorded and transcribed verbatim. Realist focus groups were conducted in Phase 1 at Site 1 by BJ, and in Phase 2 by AM and JF using realist guides (Supplementary Material 6)[28]. Yarning-style focus groups with CHW scanners were facilitated by two First Nations researchers, RC and VW[29]. Observations were documented as fieldnotes and site-visit reports by BJ, AM, JM, JY, JW, LL, AK, MB, and JF, guided by an observational template (Supplementary Material 7). Fieldnotes were recorded in secure WhatsApp groups, a low-burden tool for contemporaneous capture. Quotations were labelled by cadre and site, with brackets distinguishing participants from the same cadre and site (e.g. CHW3(2)). OFN indicated an observational field note. The Normalisation MeAsure Development (NoMAD) survey was self-administered [30] (Supplementary Material 8).

A Theory of Change, a strategic map of how the programme was expected to lead to change and how progress would be assessed, was developed during the co-design phase[13], with site-adapted versions reflecting small variations in delivery form. Theory of Change indicators were assessed jointly by BJ and AM at approximately six months (December 2023) and 24 months (July 2025) and contributed to this evaluation.

#### 2.5.2 Secondary data

Secondary data collected in Phase 3 included scan records, administrative data, and costing data. Scan records were collected and managed using REDCap V.13[31]. For Sites 2–5, data were extracted from REDCap for individuals aged 5–20 years with scans uploaded between 1 May 2024 and 30 June 2025, covering the establishment of the database to the end of Phase 2 to allow time for analysis. Analyses were restricted to participants aged 5-20, who accounted for >90% of all screening activity across the sites. This included both Stage II logbook scans and Stage III post-certification scans. At Site 1, REDCap use was limited, with most scan interpretations communicated directly by a visiting cardiologist during screening events rather than being uploaded. To estimate scanning activity, records from the cloud-based image platform, Tricefy, were used. Administrative data included site-visit dates, scanner cadres, and training progress. Data from Tricefy were also used to determine the time taken to cardiologist review. Costing data were drawn from study expenditure records, supplier and market prices, wage and workforce award data, and consultation-based estimates for timing of activities (details in Supplementary Material 9). Where remuneration ranges were available, the median rate was used for the base-case. All personnel costs incorporated a 31% on-cost.

### 2.6 Data analysis

The implementation outcomes assessment and the realist evaluation drew on the same dataset but addressed different analytical aims.

#### Implementation outcomes analysis

Implementation outcomes were reported according to the recommendations of Lengnick-Hall et al.[32], and Cronin et al.’s costing guidebook for implementation scientists[33].

Quantitative data were analysed in R (version 4.2.2). Descriptive statistics included proportions and medians with interquartile ranges (IQRs). Associations between variables were examined using Pearson correlation and simple logistic regression analyses, with results presented as correlation coefficients or odds ratios with 95% confidence intervals respectively. A significance threshold of *p* < 0.05 was applied. Fidelity for each core function was assessed using a three-point scale (‘low’, ‘medium’, ‘high’), with joint review of relevant qualitative data by BJ and AM (Table 5). Implementation strategy delivery was assessed descriptively.

**Table 5.** Fidelity rating definitions by programme component.

| Component | Fidelity measure | Low fidelity | Medium fidelity | High fidelity |
| --- | --- | --- | --- | --- |
| Community engagement | Qualitative assessment | Minimal | Intermittent | Sustained |
| Local training | % of trainees completing all training requirements | < 25% | 25–49% | ≥ 50% |
| Handheld devices | Qualitative assessment of accessibility/function | Frequent issues | Occasional issues | No issues |
| SPLASH protocol | % of uploaded scans uninterpretable<sup>1</sup> | > 40% | 11–40% | ≤ 10% |
| Image upload | Qualitative assessment of upload reliability | Frequent issues | Occasional issues | No issues |
| Cardiologist review | % of scans reviewed within 72 hours | < 50% | 50–89% | ≥ 90% |

<sup>1</sup>Threshold of ≤ 10% selected as a readily interpretable benchmark, informed by previously reported local scanner uninterpretable echocardiogram rates[22].

Cost analyses were conducted using a simplified activity-based micro-costing approach[33] and adopting a multi-provider perspective. We estimated intervention costs and implementation costs, excluding participant-borne costs, research costs, and costs beyond notifying participants of screening outcomes (Supplementary Material 9). Costed activities were identified from the fidelity assessment, non-personnel inputs were costed directly, and personnel time was estimated from fidelity data and staff validation, valued using wage rates. One-way deterministic sensitivity analyses explored uncertainty in key parameters influencing total costs. All costs were reported in 2025 Australian dollars (A$).

#### Realist evaluation analysis

Data were analysed using a realist enquiry[34, 35]. In Phase 1, BJ and AM reviewed the data and inductively grouped relevant material into ‘conceptual buckets’ representing emerging ideas. Draft context–mechanism–outcome configurations (CMOCs) were developed or refined from the initial programme theory, drawing on substantive theories where mechanisms could not be inferred directly from the data. Refined CMOCs were grouped into broader domains and synthesised narratively to produce the next iteration of the programme theory. In Phase 2, additional data were analysed deductively against existing CMOCs to assess whether they were confirmed, refuted, or required refinement. New CMOCs were developed using the same process as Phase 1, and the analysis expanded to examine subgroups. In Phase 3, quantitative data were used to explore outcome patterns and site differences.

## 3 Results

The findings draw on 36 realist interviews, 39 NoMAD surveys, and seven focus groups (totalling 26 participants), 200 researcher-days of observation across 24 months (median 5 visits per facility, range 4–9), and 360 scan records (Table 6).

**Table 6.** Overview of evaluation participants by site.

|  | Site 1 | Site 2 | Site 3 | Site 4 | Site 5 | Total |
| --- | --- | --- | --- | --- | --- | --- |
| Interviews |  |  |  |  |  | **36** |
| CHWs | 2 | 3 | 4 | 4 | 1 | 14 |
| Nurses | 1 | NA | 3 | 1 | 1 | 6 |
| GPs | NA | NA | NA | 1 | 2 | 3 |
| Managers | 2 | 2 | 2 | 2 | 1 | 9 |
| Cardiologists | NA | NA | NA | NA | NA | 4<sup>1</sup> |
| Surveys |  |  |  |  |  | **39** |
| Scanners | 7 | 3 | 5 | 4 | 4 | 24 |
| Managers | 4 | 1 | 2 | 1 | 0 | 8 |
| Other staff | 0 | 1 | 2 | 0 | 0 | 3 |
| Cardiologists | NA | NA | NA | NA | NA | 4 |
| Focus groups |  |  |  |  |  | **26** |
| Site 1 scanners |  |  |  |  |  | 5 |
| Site 1 managers |  |  |  |  |  | 3 |
| GP scanners |  |  |  |  |  | 4<sup>2</sup> |
| Nurse scanners |  |  |  |  |  | 5 |
| CHWs x 3 |  |  |  |  |  | 9 |

<sup>1</sup> Cardiologist interpretation was not site-specific as scans were pooled and reviewed off-site.
<sup>2</sup> Two additional General Practitioner (GP)–obstetricians were purposively recruited from a NEARER SCAN regional hospital study site that was demonstrating more consistent scanning than the remote sites in this evaluation.

### 3.1 Fidelity

#### Intervention fidelity

Across all sites, the *programme-in-practice* differed considerably from the *programme-as-designed* (Table 7). In total, 32 local staff commenced training (21 CHWs, 8 nurses, 3 doctors), of whom 23 (72%) were First Nations. Fourteen staff (44%) completed all training requirements during the evaluation period. Although *local training* progressed slowly (median time to certification of 14 months), a workforce capable of screening was eventually established at all sites except Site 4. When the SPLASH scans were performed, image quality was generally adequate: 87% of studies were interpretable, although the proportion of uninterpretable scans ranged from 5% to 33% across sites. Support for the programme was consistently expressed throughout the study period. However, *community engagement* ranged from sustained to minimal, which appeared to reflect the varied levels of strength and stability of relationships between the implementation team, facility staff, and the broader community.

**Table 7.** Intervention fidelity at each site.

| Site<sup>1</sup> | Community engagement | Local training | SPLASH protocol | Handheld device | Image upload |
| --- | --- | --- | --- | --- | --- |
| 1 | Medium | High | Medium | Medium | Low |
| 2 | High | High | Medium | High | Low |
| 3 | High | High | Medium | Low | Low |
| 4 | Low | Low | Medium | Low | Low |
| 5 | Low | Medium | High | Medium | Low |

<sup>1</sup>Cardiology review was not site-specific but centrally pooled and had a low fidelity overall.

Issues with the *handheld devices* were consistently reported in all sites apart from one (Site 2). This was mostly related to devices not being easily accessible or reliably charged.

> “At the end of the day you’re like ‘oh [scanning] is not that easy, someone’s taken the gel, someone’s taken the charger’…[it] ends up being a bit more challenging than **I** thought it would be” [NUR4].

Issues with image upload for off-site review were also pervasive, largely due to WiFi connectivity issues.

> “Scan done 3 days ago but hasn’t uploaded yet, still sitting in the queue, impact on turnaround time is significant” [OFN3].

*Cardiology review* of uploaded scans was slower than anticipated, with only 39% of scans reviewed within the 72-hour window, and a median time of 9 days (IQR 1–33) from upload to interpretation. A functioning operational pathway for educational feedback to scanners was not successfully established. Within the study framework, scans were reviewed by a group of cardiologists outside their usual responsibilities, with only a token payment per scan which could be optionally donated. The workflow was reported to be difficult to accommodate within usual responsibilities, particularly for lower-quality or borderline scans.

> “If they’re all normal, it’s quicker, or if the image quality is ok…it’s those borderline ones [that take time]” [CARD4].

Cardiologist rosters and email notifications when scans were uploaded were introduced as the review workflow was operationalised and refined throughout the study period. These supports were described as making reviews easier to complete within the intended timeframe.

#### Implementation strategies

Implementation strategies were delivered to differing extents across sites, with several adaptations and additional strategies introduced in response to emerging site-specific contexts. Key strategies included *ongoing training visits and screening* events (outlined in Table 2) (see timeline, Supplementary Table 1). Booster training visits, typically 2-3 days, occurred approximately twice a year and provided scanners with opportunities to refresh skills and practice with supervision. Scan activity was markedly higher on days when an expert was present (OR = 18.15, 95% CI 10.87–30.26, *p* < 0.001). Local scanners described these visits positively:

> “[Visits] definitely helped…it lifts up confidence and gets the momentum going again” [GP4].

Screening events were also conducted periodically, creating concentrated opportunities for a high volume of scans. For example, Site 3 held regular screening events, and on event days averaged 8.6 scans per site per day compared to 0.1 scans per day on non-event days. These events were commonly described as a helpful strategy for creating protected time for scanning:

> “The scheduled events are much easier for us, it frees up a bit of time” [NUR3(2)].

Conversely, uptake and perceived usefulness of some strategies varied across scanners. *Off-site support* strategies, such as WhatsApp groups or scheduled support video calls, were used by only a small number of scanners, mostly GPs or those who had strong working relationships with specific sonographers. Off-site support was perceived as having limited benefit overall:

> “I’m not a WhatsApp fan. I don’t know about the others, I don’t know why they’re not engaging” [CHW2].

Multiple strategies were introduced or adapted during implementation in response to on-the-ground contextual realities. Persistent WiFi connectivity problems prompted the provision of internet dongles, although these too had limitations:

> “The dongle does not hold charge and turns itself off” [OFN2].

Additional training supports were introduced, including *facilitating scanners to participate in a regional mass-screening week* alongside experts, and organising a community-of-practice workshop to refresh skills and share experiences. In response to persistently low scanning activity at Site 4, staff from a *second facility* serving the same community were also trained to increase screening capacity.

### 3.2 Adoption, Penetration, and Sustainability

Patterns of scanning were broadly similar across sites, with modest initial uptake and intermittent scanning activity throughout. For site-level findings see Table 8 below. 17/32 (55%) trainees performed at least one 5-20 year old logbook scan within three months of their Stage 1 training. The median time from the final day of Stage 1 training to the next logbook scan was 35 days (IQR 22–182), and the median number of scans per scanner within the first three months was 1 (IQR 1– 4). 360 scans of 5–20 year olds were performed and uploaded for off-site interpretation (either during Stage II as logbook scans or during Stage III post-certification) between May 2024 and June 2025, representing approximately 16% (360/2321) of the total 5–20 year old population across sites. Of these, 156/360 (43%) were done by CHWs. Scans were uploaded in 22% of weeks (68/305) during this period. We did not observe an increase in the frequency of scans being uploaded during the last three months.

**Table 8.** Adoption, penetration, and sustainability at each site.

|  | Site 1 | Site 2 | Site 3 | Site 4 | Site 5 |
| --- | --- | --- | --- | --- | --- |
| **Adoption<sup>1</sup>** |  |  |  |  |  |
| Trainees scanning within 3 months of commencing training (n/N, %) | 1/7 (14) | 2/2 (100) | 7/9 (78) | 6/10 (60) | 1/4 (25) |
| Median days to first scan post Stage 1 training | 163 | 33 | 32 | 5 | 321 |
| Median scans per scanner within 3 months of commencing training | 0 | 4 | 3 | 2 | 0 |
| **Penetration** |  |  |  |  |  |
| Number of scans | 63 | 31 | 120 | 20 | 126 |
| Percentage of eligible 5–20-year-olds scanned (%)<sup>2</sup> | 9 | 28 | 17 | 3 | 85 |
| Proportion of weeks in which $\geq 1$ scan was uploaded (n/N, %) | 3/61 (5) | 7/61 (11) | 20/61 (33) | 7/61 (11) | 31/61 (51) |
| **Sustainability** |  |  |  |  |  |
| Weeks with $\geq 1$ scan uploaded in final 3 months (n/N, %) | 0/13 (0) | 2/13 (15) | 3/13 (23) | 3/13 (23) | 7/13 (54) |

<sup>1</sup>Site 5 adoption figures exclude a GP who was integrated into the programme as a scanner but had echocardiography training prior to NEARER SCAN.
<sup>2</sup>No denominators are shown to preserve site anonymity. Age-specific population estimates were taken from the Australian Bureau of Statistics although these are likely an underestimate[36, 37]. For the West Pilbara site, the First Nations proportion (11.7%) was applied because the broader population is predominantly non-First Nations.

Overall, scan volumes fluctuated (Figure 3). Peaks were observed in May 2024 (during a Site 1 visit), in July 2024 (following more consistent scanning by the Site 5 GP scanner), and in March 2025 ( during a screening event at Site 3). Monthly contributions varied across sites, with Sites 3 and 5 showing relatively more consistent activity, and Sites 1, 2, and 4 showing more sporadic uploads.

**Fig. 3.**
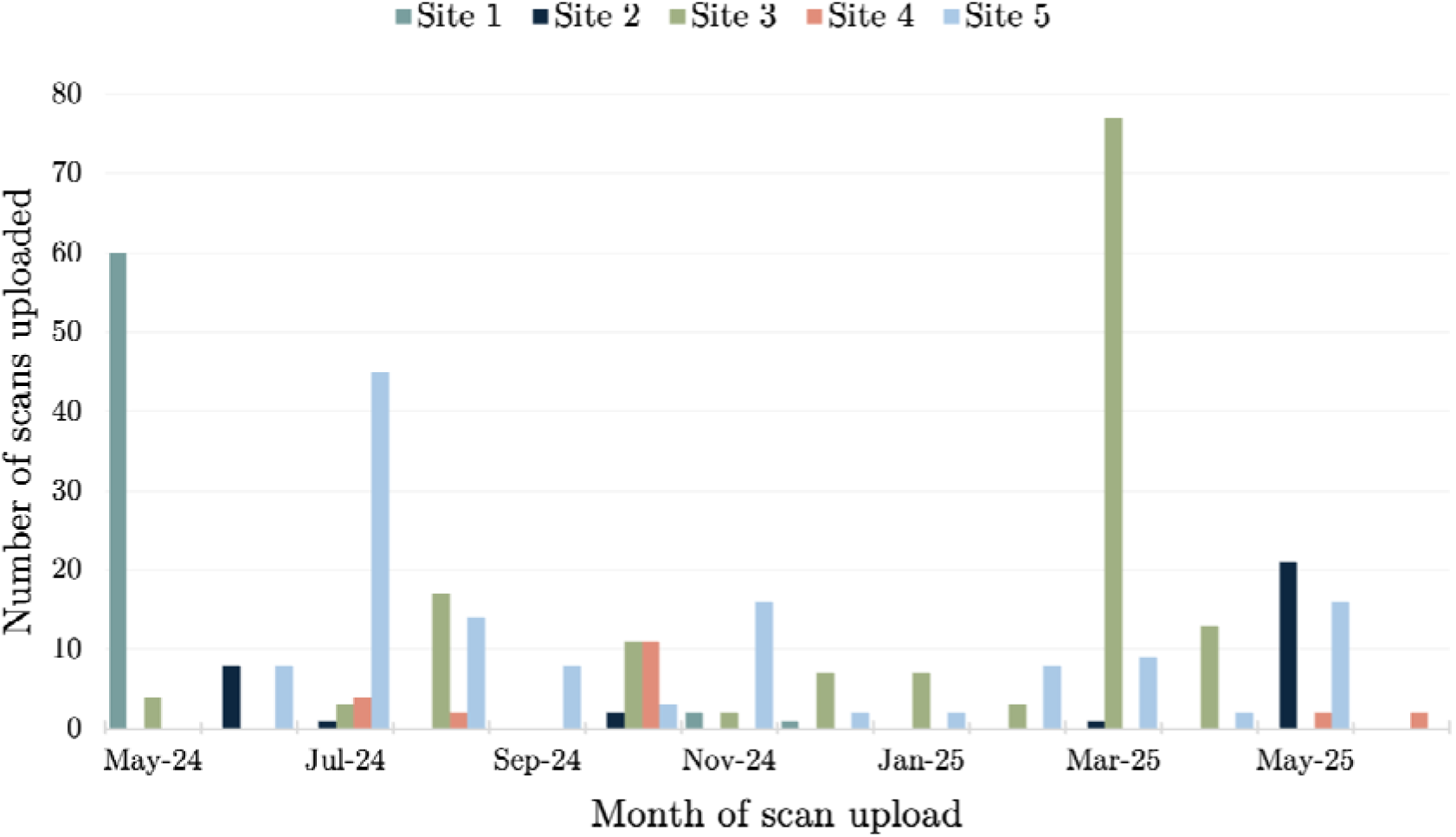
Scan activity over time by site. All months are shown, with x-axis labels displayed every second month for readability.

These findings suggested two main delivery patterns that informed the costing reported below: GP-led opportunistic screening in routine care, and CHW-led event-based screening.

### 3.3 Cost

#### Total and component costs

The estimated costs of implementing the NEARER SCAN screening programme across the participating sites are broken down into each cost component in Table 9 and Supplementary Material 9. Two screening models were costed based on the delivery patterns described above.

**Table 9.** Estimated costs of implementing the screening programme.

| Cost component | Total estimated cost | Range (dominant cost-driver) |
| --- | --- | --- |
| Training and set-up<sup>1</sup> | A\$51,093 per site | A\$46,405–A\$55,780 |
| Opportunistic screening model (GP-led) | A\$44.83 per scan | A\$35.46–A\$53.20 |
| Event-screening model (CHW-led) | A\$90.10 per scan | A\$81.09–A\$99.11 |
| Image review | A\$49.87 per scan | A\$45.78–A\$66.63 |
| Implementation package<sup>2</sup> | A\$9,858 per site per year | A\$7,067–A\$12,659 |

<sup>1</sup>The training and set-up for 32 local staff across five sites cost an estimated A\$117,193 in personnel time and A\$32,858 in support costs (including travel and accommodation), totalling A\$138,271. Up-front equipment costs (handheld devices and accessories) and equipment required for one year of scanning amounted to A\$105,413. On a per-site basis, training and set-up cost approximately A\$30,010, and equipment costs were A\$21,083, giving a total of approximately A\$51,093 per site.
<sup>2</sup>Assumes an average implementation package including two additional sonographer two-day, one-night site-visits, a recall system, promotional materials for events, audit and feedback, engagement with community organisations, graduation celebrations, and off-site assistance from sonographers.

#### Sensitivity analyses

For the training and set-up component, total costs were most sensitive to changes in personnel time and wage rates, with smaller effects from the number of CHWs trained and support costs (Figure 4).

**Fig. 4.**
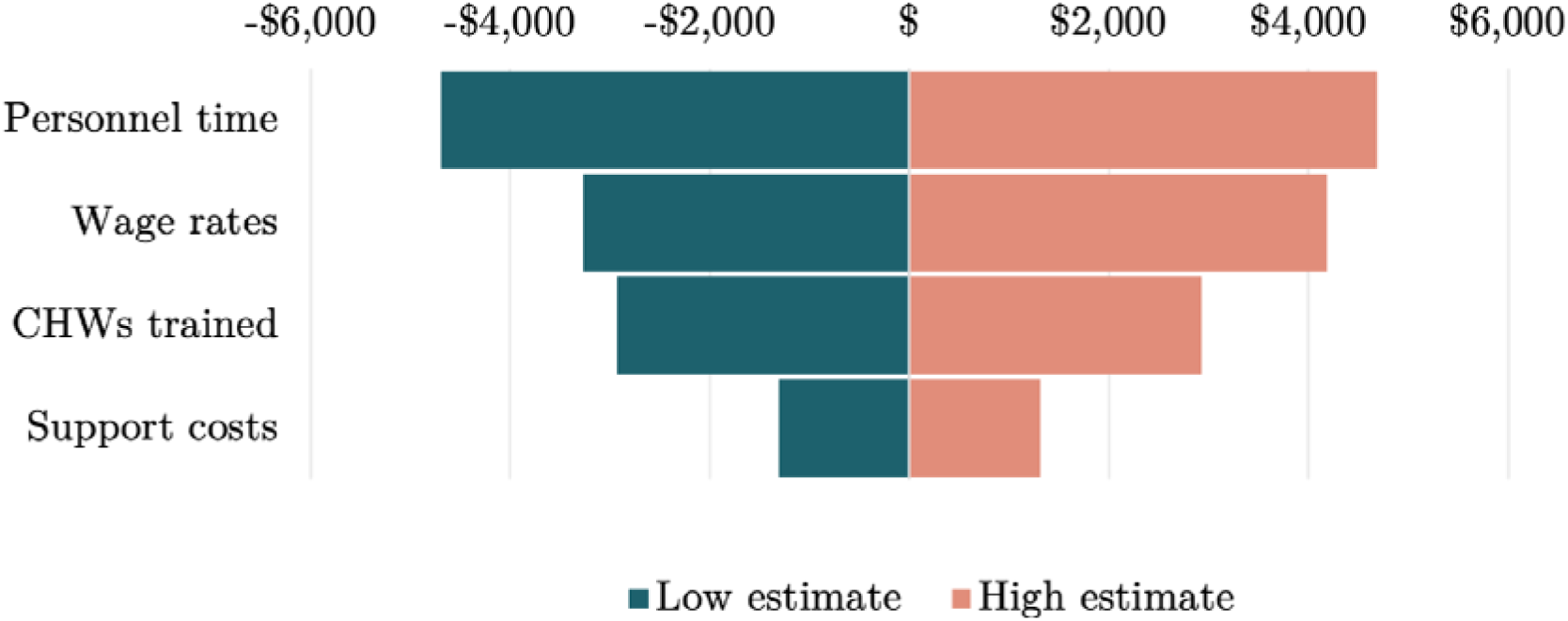
Tornado diagram showing results of the one-way deterministic sensitivity analysis for the training and set-up cost component. Bars show the change in total cost relative to the base estimate when key parameters were varied individually. Personnel time and support costs were varied by ±20%, wage rates were varied according to the minimum and maximum published award rates for each actor, and the number of CHWs trained was varied by ±2 staff from a baseline of four.

Within the screening and review components, the main driver was GP time in the opportunistic model and personnel time and support costs in the event-based model, reflecting the higher cost of sonographer time and associated travel (with smaller effects from wage variation and the number of CHWs involved) and cardiologist time and wage rates for image review. Annual implementation costs were most sensitive to the number of sonographer support visits. Detailed parameter values and tornado diagrams are provided in Supplementary Material 9.

### 3.4 Realist evaluation findings

These realist evaluation findings do not attempt to explain all observed variations in implementation but focus on CHWs and explain the conditions that limited or enabled the embedding of screening into routine work. This focus reflects that CHW-led opportunistic screening was the intended delivery approach, and is prioritised in the pilot health-system programme[8].

Acknowledging the inherent complexity, we use two overarching concepts that emerged to structure the realist findings: the legitimacy of the CHW ‘scanner role’ and the invisible work required to support opportunistic screening. The final 33 CMOCs are provided in Supplementary Table 2.

#### Legitimacy

Legitimacy is a shared perception within a group that a practice is appropriate and aligns with norms and values. The legitimacy of CHW scanning was conveyed through three forms of communication: verbal social (referring to implicit cues in everyday interactions), and written. Given their limited authority within facilities, we found that it was difficult for CHWs to generate this legitimacy on their own, and it largely depended on others.

##### Verbal communication

Managers were a key source of verbal legitimacy. When managers mentioned the programme in whole-of-staff meetings, such as the morning huddles at Site 3, the act of acknowledgement, even without explicit endorsement, shaped staff perceptions of screening as an acceptable practice (CMOC 12). A similar effect occurred in the context of managers asking the CHWs directly how scanning was going, highlighting that it was important (CMOC 14). When managers went further and actively emphasised its benefits for the community, this aligned with CHW’s professional values, and strengthened their willingness to scan (CMOC 13). Consistent with this, in the NoMAD survey, higher perceived managerial support was modestly positively correlated with perceived shared understanding of the programme: (*ρ* = 0.37, *p* = 0.032). However, support was fragile in sites that experienced frequent manager turnover where new managers often expressed support largely in principle. Without clear handover guidance this support did not translate into active communication of legitimacy (CMOC 11). Site 4, where scanning activity was lowest, experienced particularly high manager turnover.

> “I know that [the previous manager] was [supportive]. You know, really on for this but I’m not quite sure about the [new] managers, yeah, whether they will allow us that time” [CHW1(2)].

Legitimacy was communicated by staff other than managers. When colleagues referred patients to a CHW for a scan, it signalled that scanning was a recognised part of the CHW’s role (CMOC 17). This effect was particularly strong when the referral came from staff with authority, such as GPs, whose implicit endorsement through the referral carried greater weight. Referrals were more likely when scanning was visible to other staff (e.g. screening events) and when line managers explicitly supported it, as these conditions reinforced the shared expectation that scanning was a routine facility activity (CMOC 18).

> “I remember the last time [the paediatrician] was here, he called me and he asked me if I had some spare time to go with him to do a heart scan on a patient. He’s really good at doing that, very supportive. He likes to help us out to learn more about scanning” [CHW3].

##### Social communication

Visiting scanning experts, such as sonographers providing ongoing training, signalled legitimacy for prioritising screening. In remote facilities, where specialist visits are infrequent, their arrival gave staff an accepted reason to prioritise scanning while the opportunity was available (CMOC 4). However, when staff had limited opportunities to scan after the expert left, their skills declined, and some appeared to only scan during a visit (CMOC 5).

> “It was really good when [the sonographer] came back…because [when scanning] you need to lean on something” [CHW2].

Managers also communicated legitimacy through the way they organised CHW’s work. By assisting with coordinating screening events or dedicated screening clinics, they created protected time for scanning, highlighting it as a legitimate priority amidst competing demands (CMOC 8). This protected time was particularly important for CHWs and nurses, who had limited control over their own workflow and at times found it hard to independently prioritise scanning. In contrast, scanners with greater authority, such as GPs, who were generally trusted to manage their own schedules, could more readily prioritise scanning at opportune times (CMOC 20). This was reflected in scanning patterns. Among scans performed by GPs, 83% occurred when no implementation team member was on-site, compared with 14% of scans performed by CHWs, indicating GPs were more likely to scan opportunistically (OR 29.7, 95% CI 13.8–68.4, *p* < 0.001).

> “A doctor can make more independent decisions about what they do or what they need to focus on… [whereas other scanners] have people kinda of like watching those numbers [of key performance indicators]” [GP5].

When screening was conducted by more than one scanner visibly in day-to-day practice, it signalled to the wider staff team, including new staff, that scanning was a shared expectation rather than an individual initiative (CMOC 1). This was evident at Site 3, which had several scanners maintaining relatively more consistent activity, in contrast to other sites where scanning was largely carried out by one or two individuals. Beyond signalling legitimacy, having a clear nurse or GP buddy who was also trained in screening provided CHWs with support, increasing the likelihood that they scan (CMOC 3).

> “Multiple people [scanning] solidifies that this is what the health practice does, [it facilitates] more conversations about it” [GP4].

##### Written communication

Recall lists of community members eligible for screening generated through the patient record system, as used at Sites 2 and 5, signalled to both staff and patients that scanning was a legitimate activity and served as a visible prompt that made it easier to prioritise (CMOC 21). In contrast, the absence of any key performance indicator (KPI) related to scanning appeared to reduce its perceived importance for CHWs, whose workflow was often shaped by KPIs (CMOC 19).

> “To have that [recall] list and then print it, these guys [the CHWs at Site 2] are visual [learners]…‘You’re down 6 pages now guys, look at us go, do you reckon you can knock off another page?’” [CM4].
>
> “But I guess the scans aren’t…part of KPIs and…it’s not part of the managers priorities and what they’re overseeing” [FGGP5].

Implementing written legitimacy often required staff with the authority to make service-level changes. When clinical-facing staff with such authority were trained and able to engage, as with the GP scanner at Site 5, the personal investment created through being one of the scanners appeared to motivate them (CMOC 2). Similarly, when managers remained in their role long enough to build a working relationship with the implementation team, a sense of shared commitment and accountability seemed to contribute to these strategies being implemented (CMOC 10).

> “[NUR3(3)] was trying to do the [RHD screening clinic] roster” [NUR3(2)].

#### Invisible work

Invisible work refers to the under-recognised and undervalued activities generated by a practice that are nonetheless essential to making it function in everyday settings. Two forms of invisible work emerged in relation to CHWs’ efforts to embed screening. The first was logistical work: the practical tasks required to make the technology usable. The second was relational work: the cognitive and interpersonal effort involved in coordinating other priorities with colleagues and negotiating roles. Invisible work was also generated for managers and other scanners, although this is not explored here given the focus on CHWs.

##### Logistical work

How the logistical work was distributed among scanners appeared to influence scanning. For the CHWs, when these burdens were temporarily removed, such as during screening events where children were already gathered (rather than the CHW needing to locate and arrange transport of the patient), scanning shifted from feeling unmanageable to doable (CMOC 9). In addition, when the tasks required to keep scanning operational, such as charging or locating devices, were accessible to all scanners and treated as a shared expectation, this collective responsibility encouraged a more even uptake of scanning among those trained (CMOC 15). By contrast, when these tasks fell to a single scanner responsibility became concentrated and widespread use was more vulnerable to disruption.

> “We had a station for scans during [a local youth programme], like with the kids’ sports, and we were just getting two or three [scans done] at a time.” [CHW2]

Logistical work also disrupted staff expectations of how the programme should function. When routine tasks such as charging the device were not completed, scanning became harder than anticipated, leading staff to abandon intended use (CMOC 16). Recurrent upload failures also weakened confidence that scanning would lead to clinical benefit, reducing its perceived worth and undermining engagement (CMOCs 30, 33).

> “[Upload issues have] been a big disincentive because it’s been well, **I** can do the scan, but I’m gonna have to take time to try and troubleshoot to get it uploaded” [GP4].

Persistent technological issues created further invisible work that shaped engagement. Some problems became normalised, particularly those involving image upload failures, and scanners at times waited for the next expert visit to resolve them (CMOCs 31, 32). Off-site support attempts also generated invisible work that limited engagement: because CHWs were not accustomed to technology-mediated supervision, they perceived off-site troubleshooting as optional and deprioritised it amidst competing tasks (CMOC 6). Nevertheless, on the occasions when off-site support was accessed, prompt expert advice reaffirmed the clinical value of scanning and increased willingness to persist despite ongoing technical barriers (CMOC 7).

> “Scanners cannot be expected to spend the amount of time currently required to ensure images are being sent.” [OFN3]
>
> “We have really tried to make the [off-site] ‘check-ins’ work over the last year – emails, SMS, calls, and Zoom.” [OFN3]

##### Relational work

Although difficult to capture in full, CHWs alluded to the added complexity of being members of the local community. Awareness of broader family circumstances, alongside having potentially multiple overlapping responsibilities for a patient beyond physical healthcare, appeared to make it harder at times to prioritise a non-urgent task like screening. Sometimes, it was more explicit, for example when cultural protocols meant that they could not scan certain patients (CMOC 25). Further, the ‘uncovering disease’ element of screening created emotional strain at times, as the thought of detecting illness in close family members was described as difficult. Over time, these relational and emotional demands generated hidden fatigue and reduced the capacity needed to prioritise scanning that required more mental energy while still unfamiliar (CMOC 26).

> “You go to work, [and it] makes you feel extra tired, because sometimes work don’t stop, you’ve got to still worry about heart problems and stuff like that [outside of work]. Humbug [a local term that means excessive demands from family or community] after work” [CHW3(2)].
>
> “It’s tricky for [CHW4]. She’s called in lots of different directions… and then has a lot of family responsibilities on top of that. So I think for her to even have some breathing space, never mind think about doing scanning. It’s just not feasible” [GP4].

Deciding when it was appropriate to offer opportunistic screening required interpersonal judgement. CHWs and nurses reported choosing not to offer screening when it felt inappropriate, for example when a child became restless during a long appointment. They also noted that patients often declined screening when attending the facility for another specific purpose (CMOCs 22–24). Similarly, CHW and nurse scanners avoided scanning when they were unsure about local consent procedures for fear of doing the wrong thing (CMOC 29).

> “Sometimes [patients] come and they want to get their [treatment] and we are hopeful to offer a scan and they say I’ll come back tomorrow, maybe later, maybe another day” [CHW3].

Relational work appeared to vary by scanner cadre. For GPs, obtaining consent in a busy facility was often brief, as positional authority meant limited explanation was typically sufficient and patients were more likely to agree (CMOC 27). By contrast, CHWs reported providing detailed explanations, drawing on shared language and cultural proximity, and patients appeared more comfortable asking questions or seeking reassurance (CMOC 28). As a result, the same practice placed uneven relational demands, with implications for how consistently scanning could be done quickly in between other duties.

> “It is just a quick thing… I don’t think it needs a lot of explanation. If most of the times [the result] is going to be normal” [GP5]
>
> “It’s good for them to see their own people doing stuff like this, they feel comfortable…you can giggle, you can laugh, you can talk about the scan, you show them their heart” [CHW2].

## 4 Discussion

In this evaluation we found that the implementation of task-sharing echocardiographic screening with predominantly CHWs in remote First Nations communities led to identified challenges to completion of training and lower scanning activity than anticipated over the 2.5-year study period. Two broad sets of issues appeared to underpin this overall pattern. First, there were gaps in fidelity across key components of the programme: handheld devices were often not accessible or charged, image uploads were frequently unsuccessful, and off-site reviews were generally slower than planned. Second, with the exception of one GP at Site 5, opportunistic screening was rarely embedded into routine work. For the main cadre of scanners, the CHWs, this reflected two challenges: the uncertain legitimacy of their new ‘scanner’ role within busy workplaces with high staff turnover, and the extent of invisible work generated by scanning, which meant that the size of the task was greater than initially appreciated. In practice, local scanners, particularly CHWs, found it challenging to perform enough scans between sonographer visits to build their skills. As a result, certification took longer than intended, around 14 months, and scanning activity often remained low or paused entirely in the absence of in-person expert support. Activity typically increased only during expert visits or screening events.

The programme was designed largely for opportunistic screening, intended to be integrated into existing clinical encounters with eligible participants, such as child health checks. Our findings suggest that, for a CHW-led programme, under the conditions observed across sites, this model is likely to be difficult to realise sustainably. However, event-based screening emerged as a potentially effective implementation strategy to support higher screening coverage, albeit at greater cost. These findings suggest the potential for a hybrid approach, which would involve periodic screening events with visiting sonographer support, concurrent with ongoing efforts to strengthen the conditions needed for opportunistic screening between events. Recommendations to help establish these conditions are provided in Table 10. We also produced an implementation toolkit to help facilities strengthen these conditions (Supplementary Material 10).

**Table 10.** Recommendations to strengthen conditions for opportunistic screening.

| Condition | Practical actions recommended based on this evaluation |
| --- | --- |
| **Legitimacy** |  |
| Verbal communication | Managers mention scanning in staff meetings and ask scanners how scanning is going.<br>Encourage other staff to refer eligible patients for scanning. |
| Social communication | Create protected time through screening events or dedicated scanning clinics.<br>Provide multiple support visits from sonographers each year.<br>Train multiple CHW scanners and a senior nurse or GP. |
| Written communication | Create recall lists of eligible participants that require a scan.<br>Include scanning in job descriptions and KPIs.<br>Develop a scanning handover plan for incoming managers in the event of staff turnover. |
| **Invisible work** |  |
| Logistical work | Provide internet dongles for periods of unstable internet connection for uploading scans.<br>Identify a secure location to store and charge devices that is accessible to all scanners. |
| Relational work | Acknowledge that local staff may need more time for screening interactions.<br>Develop local consent processes that all scanners are comfortable using.<br>Confirm local cultural considerations with local staff before implementing screening. |

The appropriate balance between opportunistic and event-based screening will need to be determined by individual facilities and communities, guided by local priorities and workforce capacity. For example, in smaller communities, fewer eligible participants could spread scanning opportunities thinly in an opportunistic model, making skill consolidation harder than when scans are clustered at occasional high-volume events. The preliminary costing reported provides an initial indication of implementation resource requirements and highlights that sustaining the programme will require funding for ongoing implementation, not only initial training.

A similar task-shifting echocardiographic screening programme for RHD, in which local scanners acquire and interpret images, is being implemented through the ADUNU programme in northern Uganda[38, 39]. In its first year, ADUNU screened more than 11,000 people through facility-based screening and outreach events, supporting the feasibility of these novel workforce approaches for early detection at scale[38]. However, progression from initial training to certification was also challenging within the ADUNU programme, with only 17 of 58 Stage I trainees completing Stage II and gaining certification in the first year[38].

The implementation difficulties we observed are likely not unique to RHD screening but reflect broader challenges in sustaining screening in remote First Nations communities. Australia’s national diabetic retinopathy screening programme uses a similar task-sharing model, introducing imaging technology into remote facilities and, in some settings, training staff to opportunistically acquire retinal photographs for off-site review [40, 41]. Despite being a more mature, nationally funded programme, only 19/132 (14%) sites uploaded images over a nine-month period, and in the Northern Territory, only 79 First Nations people were screened in 2022–23 despite an estimated 3,477 eligible people living with diabetes[42–44].

Our findings suggest that establishing the legitimacy of a new role is a dynamic, negotiated process that is continually reassessed[45–47]. Low role clarity, combined with limited authority, high workload, and shifting norms in high-turnover teams, can constrain embedding by leaving little scope for CHWs to actively negotiate the legitimacy of the role. In our evaluation, CHWs’ ‘scanner’ role tended to emerge more passively and was shaped largely by others. Reay and colleagues showed that nursing leaders cultivated opportunities to legitimise a new nurse practitioner role by drawing on social connections and system knowledge[48]. Our findings suggest negotiating legitimacy is likely conditional on an actors’ position within workplace hierarchies. We also found that when a new technology introduces an entirely new role, legitimacy and use reinforce one another[49]. Our study adds a temporal dimension by suggesting that during early embedding, legitimacy appears to shape use more strongly than use builds legitimacy, and therefore warrants explicit attention from the outset of implementation.

This evaluation also demonstrates that the support for invisible work shapes embedding. Building on Star and Strauss’ description of invisible work as essential[50], we suggest it is not only the presence of this work that matters, but how it is distributed and coordinated. When logistical tasks were shared and predictable, they reinforced collective responsibility. The invisible relational work of First Nations CHWs highlighted in our evaluation echoes that described by Topp and colleagues[17, 18, 51] and provides an empirical example of how this relational work can both enable and constrain embedding.

### Strengths and limitations

This is among the earliest implementation science evaluations of echocardiographic RHD screening and a task-sharing programme involving First Nations CHWs in Australia. The prospective, mixed-methods, multi-site design enabled us to follow implementation outcomes over time (including understudied outcomes such as cost) and to triangulate quantitative outcomes data with qualitative accounts of contextual factors and mechanisms, supporting a more explanatory understanding of the conditions required to support screening. Without this evaluation, there would be a greater risk of Type III error, whereby limited impact might be attributed to the screening programme itself rather than to challenges in its implementation [52].

Several limitations may have influenced results. Fidelity ratings were based on evaluator judgement, albeit informed in some cases by quantitative measures, and are therefore subject to interpretation. Scan records may have included repeat scans for the same participant, which could overestimate scan counts and apparent coverage. Our assessment of sustainability reflects only early maintenance of scanning activity over a relatively short period and does not capture longer-term sustainability. The costing analysis was deliberately simple. The perspectives captured were of healthcare staff and we did not collect data directly from screening participants. Finally, the mechanisms we describe relate specifically to First Nations CHWs in Australia. While concepts such as legitimacy and invisible work are likely to be relevant for other cadres and settings, especially those who occupy similar positions within workplace hierarchies, different contextual factors may be more influential elsewhere, and transferability should therefore be judged with careful attention to local context.

Future work should examine the governance, policy, and funding arrangements needed to support implementation and longer-term sustainability during scale-up. Costing should be extended to a cost-effectiveness analyses.

## 5 Conclusion

Implementation of task-sharing echocardiographic screening for RHD in remote First Nations communities led to a lower and more variable scanning frequency than anticipated with fidelity gaps across key programme components and limited embedding of scanning in CHWs’ routine work. The realist evaluation showed that progress towards routine scanning for CHWs hinged on the legitimacy of their role as a scanner and on recognition and support of the invisible logistical and relational work that scanning entails. Despite these challenges, ongoing community support for RHD screening with its foundations in local capacity building, suggest that continued refinement is warranted. Overall, the findings support a largely event-based model in which periodic, well-supported screening events form the core mode of delivery, while facilities incrementally strengthen the conditions for CHW-led opportunistic scanning between events.

## Data Availability

The datasets generated and analysed during the current study are not publicly available due to risk of identification of participants but a summary will be available from the corresponding author on reasonable request.

## Acknowledgements

We thank the First Nations communities, Elders, and all healthcare staff who contributed their time, knowledge, and guidance to this work. We also acknowledge the support of local and regional health authorities, schools, and other community organisations who helped to enable the study. We are also grateful to Karla Canuto, Bart Currie, Marisa Gilles, Paul Burgess, David Simon, Jess De Dassel, Peter Morris, and Holger Unger for their support as investigators on the NEARER SCAN grant. We further thank the cardiologists who generously gave their time to review scans.

**Trial registration:** ClinicalTrials.gov identifier NCT06002243, registered 21 August 2023.

## Declarations

Ethics approval was obtained by the Human Research Ethics Committee of Northern Territory Health and Menzies School of Health Research (Reference Number: 2022-4479), the WA Aboriginal Health Ethics Committee (References Number: HREC1237), and the University of Oxford’s Tropical Research Ethics Committee (Reference Number: 519-23). The study was conducted in accordance with the Declaration of Helsinki.

## Contributors

B.J. took principal responsibility for leading the study, including study design, data collection, analysis, interpretation, and manuscript preparation. M.E., J.F., and S.N. provided senior supervision and oversight of the study, and J.C. supervised the costing component. A.M. contributed to data collection, analysis, and interpretation. J.M. and J.Y. co-led the programme of work and contributed to data collection and interpretation. J.W., L.L., and A.K. contributed to data collection and provided expert input on echocardiographic components. M.B. coordinated the project and contributed to costing data collection. R.C., L.M., V.W., and D.F. contributed to data collection. A.P. contributed to data interpretation and manuscript preparation. B.R., A.P.R., G.W., E.H., J.M.K., N.J.H., P.R., A.F.M., H.H., J.O., G.S.H., D.E., A.B., and A.C.S. provided expert input and contributed to interpretation of findings. K.B., J.G., S.L., G.P., M.S., and B.S. were key local stakeholders and provided community-based expertise. All authors reviewed and approved the final manuscript. The following authors had access to the full dataset and verified the data: B.J., A.M., and J.F.

## Data sharing agreement

The datasets generated and analysed in this study are not publicly available due to risk of identification of participants but a summary will be available from the corresponding author on reasonable request. The study protocol is available [16], and details of the co-design process are described elsewhere [13].

## Footnotes

1 *We use ‘community health worker’ as a collective term for an Aboriginal and/or Torres Strait Islander (a) health practitioner, (b) health worker, or (c) community worker employed by a health facility without formal health qualifications. These cadres are similar to that of community health workers internationally*.

## Notes

### Competing Interest Statement

The authors have declared no competing interest.

### Clinical Protocols

https://bmjopen.bmj.com/content/14/10/e083467.abstract

### Author Declarations

Ethics approval was obtained by the Human Research Ethics Committee of Northern Territory Health and Menzies School of Health Research (Reference Number: 2022-4479), the WA Aboriginal Health Ethics Committee (References Number: HREC1237), and the University of Oxfords Tropical Research Ethics Committee (Reference Number: 519-23). The study was conducted in accordance with the Declaration of Helsinki.

