## Supplementary Material for "Task-sharing echocardiographic screening for rheumatic heart disease in remote First Nations Australian communities: implementation evaluation from the NEARER SCAN study"

**Supplementary Online Content**

Supplementary Material 1. StaRI statement

Supplementary Material 2. RAMESE II statement

Supplementary Material 3. Initial Programme Theory

Supplementary Material 4. TIDieR checklist

Supplementary Material 5. Example interview guide

Supplementary Material 6. Example focus group guide

Supplementary Material 7. Observational fieldnote template

Supplementary Material 8. NoMAD survey

Supplementary Material 9. Costing workbook

Supplementary Table 1. Training and supervision dose by site

Supplementary Table 2. Finals CMOCs

Supplementary Material 10. Implementation toolkit

Supplementary Material 1. StaRI statement

#### Standards for Reporting Implementation Studies: the StaRI checklist for completion

The StaRI standard should be referenced as: Pinnock H, Barwick M, Carpenter C, Eldridge S, Grandes G, Griffiths CJ, Rycroft-Malone J, Meissner P, Murray E, Patel A, Sheikh A, Taylor SJC for the StaRI Group. Standards for Reporting Implementation Studies (StaRI) statement. *BMJ* 2017;356:i6795

The detailed Explanation and Elaboration document, which provides the rationale and exemplar text for all these items is: Pinnock H, Barwick M, Carpenter C, Eldridge S, Grandes G, Griffiths C, Rycroft-Malone J, Meissner P, Murray E, Patel A, Sheikh A, Taylor S, for the StaRI group. Standards for Reporting Implementation Studies (StaRI). Explanation and Elaboration document. *BMJ Open* 2017 2017;7:e013318

Notes: A key concept of the StaRI standards is the dual strands of describing, on the one hand, the implementation strategy and, on the other, the clinical, healthcare, or public health intervention that is being implemented. These strands are represented as two columns in the checklist.

| | |
|---|---|
| The primary focus of implementation science is the implementation strategy (column 1) and the expectation is that this will always be completed. | The evidence about the impact of the intervention on the targeted population should always be considered (column 2) and either health outcomes reported or robust evidence cited to support a known beneficial effect of the intervention on the health of individuals or populations. |

The StaRI standardsrefers to the broad range of study designs employed in implementation science. Authors should refer to other reporting standards for advice on reporting specific methodological features. Conversely, whilst all items are worthy of consideration, not all items will be applicable to, or feasible within every study.

| Checklist item | | Reported on page # | Implementation Strategy | Reported on page # | Intervention |
|---|---|---|---|---|---|
| | | | "Implementation strategy" refers to how the intervention was implemented | | "Intervention" refers to the healthcare or public health intervention that is being implemented. |
| **Title and abstract** | | | | | |
| Title | **1** | 1 | Identification as an implementation study, and description of the methodology in the title and/or keywords | | |
| Abstract | **2** | 2 | Identification as an implementation study, including a description of the implementation strategy to be tested, the evidence-based intervention being implemented, and defining the key implementation and health outcomes. | | |

| | | | | | |
|---|---|---|---|---|---|
| **Introduction** | | | | | |
| Introduction | **3** | 3 | Description of the problem, challenge or deficiency in healthcare or public health that the intervention being implemented aims to address. | | |
| Rationale | **4** | 4 and previously published | The scientific background and rationale for the implementation strategy (including any underpinning theory/framework/model, how it is expected to achieve its effects and any pilot work). | 6 and previously published | The scientific background and rationale for the intervention being implemented (including evidence about its effectiveness and how it is expected to achieve its effects). |
| Aims and objectives | **5** | 3 | The aims of the study, differentiating between implementation objectives and any intervention objectives. | | |
| **Methods: description** | | | | | |
| Design | **6** | 4 | The design and key features of the evaluation, (cross referencing to any appropriate methodology reporting standards) and any changes to study protocol, with reasons | | |
| Context | **7** | 5 | The context in which the intervention was implemented. (Consider social, economic, policy, healthcare, organisational barriers and facilitators that might influence implementation elsewhere). | | |
| Targeted 'sites' | **8** | 5 | The characteristics of the targeted 'site(s)' (e.g locations/personnel/resources etc.) for implementation and any eligibility criteria. | 5,6 | The population targeted by the intervention and any eligibility criteria. |
| Description | **9** | 7 | A description of the implementation strategy | 5,6 | A description of the intervention |
| Sub-groups | **10** | 7,8,9 | Any sub-groups recruited for additional research tasks, and/or nested studies are described | | |
| **Methods: evaluation** | | | | | |
| Outcomes | **11** | 9,10 | Defined pre-specified primary and other outcome(s) of the implementation strategy, and how they were assessed. Document any pre-determined targets | Not assessed here, will be | Defined pre-specified primary and other outcome(s) of the intervention (if assessed), and how they were assessed. Document any pre-determined targets |

| | | | | | |
|---|---|---|---|---|---|
| | | | | reported separately | |
| Process evaluation | **12** | 10,11 | Process evaluation objectives and outcomes related to the mechanism by which the strategy is expected to work | | |
| Economic evaluation | **13** | 10 | Methods for resource use, costs, economic outcomes and analysis for the implementation strategy | 10 | Methods for resource use, costs, economic outcomes and analysis for the intervention |
| Sample size | **14** | Not reported (quant), qualitative sampling described 9–10 | Rationale for sample sizes (including sample size calculations, budgetary constraints, practical considerations, data saturation, as appropriate) | | |
| Analysis | **15** | 9,10,11 | Methods of analysis (with reasons for that choice) | | |
| Sub-group analyses | **16** | N/A | Any a priori sub-group analyses (e.g. between different sites in a multicentre study, different clinical or demographic populations), and sub-groups recruited to specific nested research tasks | | |

| **Results** | | | | | |
|---|---|---|---|---|---|
| Characteristics | **17** | 15 | Proportion recruited and characteristics of the recipient population for the implementation strategy | 15 | Proportion recruited and characteristics (if appropriate) of the recipient population for the intervention |
| Outcomes | **18** | 15 | Primary and other outcome(s) of the implementation strategy | - | Primary and other outcome(s) of the Intervention (if assessed) |
| Process outcomes | **19** | 12 | Process data related to the implementation strategy mapped to the mechanism by which the strategy is expected to work | | |

| | | | | | |
|---|---|---|---|---|---|
| Economic evaluation | **20** | 16, 17 | Resource use, costs, economic outcomes and analysis for the implementation strategy | 16, 17 | Resource use, costs, economic outcomes and analysis for the intervention |
| Sub-group analyses | **21** | 16–17 (site comparisons) | Representativeness and outcomes of subgroups including those recruited to specific research tasks | | |
| Fidelity/ adaptation | **22** | 12,13,14 | Fidelity to implementation strategy as planned and adaptation to suit context and preferences | 12,13,14 | Fidelity to delivering the core components of intervention (where measured) |
| Contextual changes | **23** | 12,13,14 | Contextual changes (if any) which may have affected outcomes | | |
| Harms | **24** | No formal harms reported, emotional burden described 19-22 | All important harms or unintended effects in each group | | |
| **Discussion** | | | | | |
| Structured discussion | **25** | 22-25 | Summary of findings, strengths and limitations, comparisons with other studies, conclusions and implications | | |
| Implications | **26** | 22-25 | Discussion of policy, practice and/or research implications of the implementation strategy (specifically including scalability) | 22-25 | Discussion of policy, practice and/or research implications of the intervention (specifically including sustainability) |
| **General** | | | | | |
| Statements | **27** | 26 | Include statement(s) on regulatory approvals (including, as appropriate, ethical approval, confidential use of routine data, governance approval), trial/study registration (availability of protocol), funding and conflicts of interest | | |

Supplementary Materials 2. RAMESE II statement

| | | **Reported in document [Y/N/Unclear]** | **Page(s) in document** |
|---|---|---|---|
| | TITLE | | |
| 1 | In the title, identify the document as a realist evaluation | Y | pp. 1 |
| | SUMMARY OR ABSTRACT | | |
| 2 | Journal articles will usually require an abstract, while reports and other forms of publication will usually benefit from a short summary. The abstract or summary should include brief details on: the policy, programme or initiative under evaluation; programme setting; purpose of the evaluation; evaluation question(s) and/or objective(s); evaluation strategy; data collection, documentation and analysis methods; key findings and conclusions Where journals require it and the nature of the study is appropriate, brief details of respondents to the evaluation and recruitment and sampling processes may also be included. Sufficient detail should be provided to identify that a realist approach was used and that realist programme theory was developed and/or refined | Y | pp. 2 |
| | INTRODUCTION | | |
| 3 | *Rationale for evaluation*<br>Explain the purpose of the evaluation and the implications for its focus and design | Y | pp. 3 |
| 4 | *Programme theory*<br>Describe the initial programme theory (or theories) that underpin the programme, policy or initiative | Y | pp. previously reported, and also reported in additional file 3 |
| 5 | *Evaluation questions, objectives and focus*<br>State the evaluation question(s) and specify the objectives for the evaluation. Describe whether and how the programme theory was used to define the scope and focus of the evaluation | Y | pp. 3 |
| 6 | *Ethical approval*<br>State whether the realist evaluation required and has gained ethical approval from the relevant authorities, providing details as appropriate. If ethical approval was deemed unnecessary, explain why | Y | pp. 26 |
| | METHODS | | |
| 7 | *Rationale for using realist evaluation*<br>Explain why a realist evaluation approach was chosen and (if relevant) adapted | Y | pp. 4 |
| 8 | *Environment surrounding the evaluation*<br>Describe the environment in which the evaluation took place | Y | pp. 5 |
| 9 | Describe the programme policy, initiative or product evaluated | Y | pp. 5 |

| | | | |
|---|---|---|---|
| | Provide relevant details on the programme, policy or initiative evaluated | | |
| 10 | *Describe and justify the evaluation design*<br>A description and justification of the evaluation design (i.e. the account of what was planned, done and why) should be included, at least in summary form or as an appendix, in the document which presents the main findings. If this is not done, the omission should be justified and a reference or link to the evaluation design given. It may also be useful to publish or make freely available (e.g. online on a website) any original evaluation design document or protocol, where they exist | Y | pp. 4 |
| 11 | *Data collection methods*<br>Describe and justify the data collection methods – which ones were used, why and how they fed into developing, supporting, refuting or refining programme theory<br>Provide details of the steps taken to enhance the trustworthiness of data collection and documentation | Y | pp. 7 |
| 12 | *Recruitment process and sampling strategy*<br>Describe how respondents to the evaluation were recruited or engaged and how the sample contributed to the development, support, refutation or refinement of programme theory | Y | pp. 8 |
| 13 | *Data analysis*<br>Describe in detail how data were analysed. This section should include information on the constructs that were identified, the process of analysis, how the programme theory was further developed, supported, refuted and refined, and (where relevant) how analysis changed as the evaluation unfolded | Y | pp. 9 |
| | RESULTS | | |
| 14 | *Details of participants*<br>Report (if applicable) who took part in the evaluation, the details of the data they provided and how the data was used to develop, support, refute or refine programme theory | Y | pp. 12 |
| 15 | *Main findings*<br>Present the key findings, linking them to contexts, mechanisms and outcome configurations. Show how they were used to further develop, test or refine the programme theory | Y | pp. 18 |
| | DISCUSSION | | |
| 16 | *Summary of findings*<br>Summarise the main findings with attention to the evaluation questions, purpose of the evaluation, programme theory and intended audience | Y | pp. 22 |
| 17 | *Strengths, limitations and future directions* | Y | pp. 24 |

| | | | |
|---|---|---|---|
| | Discuss both the strengths of the evaluation and its limitations. These should include (but need not be limited to): (1) consideration of all the steps in the evaluation processes; and (2) comment on the adequacy, trustworthiness and value of the explanatory insights which emerged<br>In many evaluations, there will be an expectation to provide guidance on future directions for the programme, policy or initiative, its implementation and/or design. The particular implications arising from the realist nature of the findings should be reflected in these discussions | | |
| 18 | *Comparison with existing literature*<br>Where appropriate, compare and contrast the evaluation's findings with the existing literature on similar programmes, policies or initiatives | Y | pp. 16-18 |
| 19 | *Conclusion and recommendations*<br>List the main conclusions that are justified by the analyses of the data. If appropriate, oder recommendations consistent with a realist approach | Y | pp. 19 |
| 20 | *Funding and conflict of interest*<br>State the funding source (if any) for the evaluation, the role played by the funder (if any) and any conflicts of interests of the evaluators | Y | pp. 19 |

Source: Wong, G., Westhorp, G., Manzano, A., Greenhalgh, J., Jagosh, J., & Greenhalgh, T. (2016). RAMESES II reporting standards for realist evaluations. *BMC medicine*, *14*(1), 96. https://doi.org/10.1186/s12916-016-0643-1

Supplementary Material 3. Initial Programme Theory

The initial programme theory proposes that implementation of the programme into routine practice in remote First Nations communities can be supported if the clinical role of the scanners is protected, and the programme logistics are flexible.

Protecting the clinical role of scanners begins with how the programme is introduced to a clinic. In smaller facilities, versatile roles, close working relationships, and overlapping duties amplify each staff member's influence on programme integration. Consequently, staff awareness and perceptions of the scanners' clinical role may shape implementation (IPT 1). Identifying informal leaders who influence team attitudes and ensuring they support the programme may promote its acceptance (IPT 2).

When presenting the programme, it is important to consider which aspects to emphasise. If scanners are professionally disempowered, as FNCHWs in these settings may be, highlighting the programme's upskilling potential may trigger a collective sense of responsibility for capacity building (IPT 3). If staff hold concerns about scanners' clinical competence, emphasising the evidence supporting the model's safety, for example, diagnostic accuracy studies or the World Heart Federation guidelines, may help build confidence and support (IPT 4).

After the programme is introduced, training of local health staff begins. When deciding who to train, consideration should be given to how best to sustain scanning in the facility. One strategy may be to train multiple staff members, so that scanners have immediate access to trusted, on-site support when technical or procedural issues arise (IPT 5). In smaller communities where screening volumes are expected to be lower, maintaining competence and confidence may be more challenging. While training multiple staff members can provide immediate collegial support, it may also dilute practice opportunities and hinder skill consolidation. It is also important to prioritise training staff from the local community, who are more likely to remain in the role long term because of their connection to the place and people (IPT 6).

From a logistical perspective, low-dose, high-frequency in-person training may suit the constraints of busy remote facilities and help maintain scanners' self-belief and technical competence over time (IPT 7). Furthermore, fostering a sense of supported ownership over the clinical role could encourage scanners to undertake programme-adjacent activities, such as restocking gel, charging devices, or addressing software and hardware issues (IPT 8).

Following training, local health staff begin performing scans independently. Early success is often pivotal, as initial failures may generate doubts about the workability of the programme and lead to abandonment or deprioritisation of the task in favour of more familiar routines (IPT 9). Empowering scanners to adapt the programme's logistics to fit their workflow might help mitigate early implementation challenges (IPT 10).

The transition to the new role could also be supported by establishing formal protocols and ensuring staff are allocated sufficient time to perform scans. Such protocols may serve as an informal endorsement and form of permission for trained staff (IPT 11), while protected time signals institutional valuing of the task within daily practice (IPT 12).

The technical and specialised nature of scanning may enhance job satisfaction for those trained, owing to increased respect from colleagues and patients (IPT 13). However, this same interest may create vulnerability if new or transient staff (e.g. rotating GPs) assume responsibility for scanning, disrupting the developing role integration. When these staff depart, the facility may be left without anyone confident to continue scanning (IPT 14), a particular concern given the high turnover typical of remote Australian facilities.

The off-site review of images by experts may also shape programme integration. Constructive feedback from reviewers could help sustain the motivation of scanners (IPT 15). Establishing formal governance structures to coordinate this interaction may further strengthen accountability and consistency. However, because tasks aligned with day-to-day responsibilities are often prioritised over more distant, ad-hoc tasks, it is important that expert reviewers are clearly informed of their expected turnaround times. Delays in feedback may discourage scanners, who could become disheartened and less inclined to continue scanning (IPT 16).

Overall, this initial programme theory served as a starting point for the empirical testing and refinement of the theory.

Supplementary Material 4. TIDieR checklist

TIDieR
Template for Intervention Description and Replication

#### The TIDieR (Template for Intervention Description and Replication) Checklist*:

Information to include when describing an intervention and the location of the information

| Item number | Item | Where located ** Primary paper (page or appendix number) | Other † (details) |
|---|---|---|---|
| | **BRIEF NAME** | | |
| **1.** | Provide the name or a phrase that describes the intervention. | 1 | ______________ |
| | **WHY** | | |
| **2.** | Describe any rationale, theory, or goal of the elements essential to the intervention. | 3,5,6 | Protocol and co-design papers cited in text |
| | **WHAT** | | |
| **3.** | Materials: Describe any physical or informational materials used in the intervention, including those provided to participants or used in intervention delivery or in training of intervention providers. Provide information on where the materials can be accessed (e.g. online appendix, URL). | 5,6 | ______________ _ |
| **4.** | Procedures: Describe each of the procedures, activities, and/or processes used in the intervention, including any enabling or support activities. | 5,6 | ______________ _ |

| | | | |
|---|---|---|---|
| | **WHO PROVIDED** | | |
| **5.** | For each category of intervention provider (e.g. psychologist, nursing assistant), describe their expertise, background and any specific training given. | 3,5,6,7 | ______________ |
| | **HOW** | | |
| **6.** | Describe the modes of delivery (e.g. face-to-face or by some other mechanism, such as internet or telephone) of the intervention and whether it was provided individually or in a group. | 3,5,6,7 | ______________ |
| | **WHERE** | | |
| **7.** | Describe the type(s) of location(s) where the intervention occurred, including any necessary infrastructure or relevant features. | 5,6 | ______________ |
| | **WHEN and HOW MUCH** | | |
| **8.** | Describe the number of times the intervention was delivered and over what period of time including the number of sessions, their schedule, and their duration, intensity or dose. | 4,6,12,13 | |
| | **TAILORING** | | |
| **9.** | If the intervention was planned to be personalised, titrated or adapted, then describe what, why, when, and how. | 4,7 | ______________ |
| | **MODIFICATIONS** | | |
| **10.**‡ | If the intervention was modified during the course of the study, describe the changes (what, why, when, and how). | 12,13 | ______________ |
| | **HOW WELL** | | |

| | | | |
|---|---|---|---|
| **11.** | Planned: If intervention adherence or fidelity was assessed, describe how and by whom, and if any strategies were used to maintain or improve fidelity, describe them. | 9,10 | ______________ _ |
| **12.‡** | Actual: If intervention adherence or fidelity was assessed, describe the extent to which the intervention was delivered as planned. | 11,12,13,14 | ______________ _ |

** **Authors** - use N/A if an item is not applicable for the intervention being described. **Reviewers** – use '?' if information about the element is not reported/not sufficiently reported.

† If the information is not provided in the primary paper, give details of where this information is available. This may include locations such as a published protocol or other published papers (provide citation details) or a website (provide the URL).

‡ If completing the TIDieR checklist for a protocol, these items are not relevant to the protocol and cannot be described until the study is complete.

* We strongly recommend using this checklist in conjunction with the TIDieR guide (see *BMJ* 2014;348:g1687) which contains an explanation and elaboration for each item.

* The focus of TIDieR is on reporting details of the intervention elements (and where relevant, comparison elements) of a study. Other elements and methodological features of studies are covered by other reporting statements and checklists and have not been duplicated as part of the TIDieR checklist. When a **randomised trial** is being reported, the TIDieR checklist should be used in conjunction with the CONSORT statement (see www.consort-statement.org) as an extension of **Item 5 of the CONSORT 2010 Statement.** When a **clinical trial protocol** is being reported, the TIDieR checklist should be used in conjunction with the SPIRIT statement as an extension of **Item 11 of the SPIRIT 2013 Statement** (see www.spirit-statement.org). For alternate study designs, TIDieR can be used in conjunction with the appropriate checklist for that study design (see www.equator-network.org).

Supplementary Material 5. Example interview guide

Phase 2 interview guide for CHWs at Site 4

Thank you for your time. As you know we have been evaluating the NEARER SCAN programme to understand best ways to integrate it into usual care. We are specifically interested in finding out across the sites, what works, for whom, when, how, and why. If we can answer these questions, we can integrate the programme more successfully for more sites using what we have found.

I was hoping to put a few suggestions of what we have found so far to you today and get your feedback. I would really love to hear what you think of these and encourage you to respond and challenge based on your experience, and by providing examples or counterexamples.

It would not be helpful for us as an outcome of this evaluation to point out the hurdles to implementing the programme; you and we already know these (e.g. staff turnover, no time, limited resources). What we are interested in is: given these are present all the time, how come sometimes they stop people from scanning but other times people manage to scan despite them?

Questions:

- We know that it has been quite hard for you to scan here in [Site 4]. We think that this might be because you have not had much support from the managers to scan. Do you think that is the case?
- In other sites, some things that have helped are when scanner colleagues refer patients for them to scan. Does this ever happen here?
- Do you think some of the things that have made it harder to scan here are not just about NEARER SCAN but about the clinic in general?
- Are you happy to talk about some of those issues?
- In some clinics the manager is the same for a long period, in others the manager and staff come and go. We think this makes it harder to integrate new programmes. What do you think about that?
- We saw that most people scanned seemed to be interested in learning more about their heart, for example the valves and RHD. Was this the case for the people you scanned? Were you able to have these discussions at the time?

Supplementary Material 6. Example focus group guide

Phase 2 focus group guide for doctors across sites

Tips for 'realist' interviewing}

- At this stage, as the evaluation nears completion, focus on what has actually occurred in response to the statements read aloud, rather than what participants hope to see happen.
- Keep the discussion focused on reasons behind successful/unsuccessful implementation, rather than general thoughts and experiences.
- Use "what makes" or "what happens when" questions to elicit explanations (e.g. What makes you feel hesitant to scan?").
- Use "what if" hypotheticals to explore variations (e.g."What if you couldn't access the scanner, what would you do?").

Questions:

- For GPOa and GPOb: Can you describe your experience with implementing NEARER SCAN in antenatal care at this hospital?
- For all: Compared to when NEARER SCAN first started, what changes (if any) have you noticed in how scanning fits into your routine care?
- For GPOa and GPOb: What do you think has contributed most to the success of NEARER SCAN at your site?
- For GPOa and GPOb: One of the things we have found so far in the remote sites is that even though we had envisioned scanning to be done opportunistically, there are many more scans performed during scheduled events. How have you found scanning opportunistically and tagging onto your antenatal check-ups? Why did this work?
- For GP4 and GP5: How do you find scanning opportunistically rather than scheduled screening? If easier for you than the FNCHWs, why?

Factors:

When GPs build momentum in their scanning, a burst in scanning experience builds their confidence regardless of whether they have completed training, but confidence alone does not ensure they will continue scanning in practice.

When  did you start scanning more frequently? What was the relationship to finishing the 100 training scans and being assessed and passed as competent?

Patients are more likely to accept and complete an opportunistic scan when offered by a doctor rather than by a CHW or nurse, due to the perceived medical authority of doctors and implicit power dynamics.

- How do patients typically respond when you offer an echocardiographic scan during an antenatal visit?
- Some evidence suggests that patients feel a greater obligation to accept a scan when offered by a doctor, does this align with your experience?
- Are there situations where patients are more or less likely to accept an opportunistic scan? What influences this decision?

When scanning administration (e.g.device maintenance, uploads, troubleshooting) is not explicitly assigned, it falls to staff perceived as having more time, tech skills, or English proficiency, leading to frustration and workload imbalance.

- For GPOa and GPOb: In your experience, how has scanning-related administration (e.g. uploading images, device maintenance) been distributed among staff here?
- Do you feel that certain staff members have taken on a disproportionate amount of the administrative burden? If so, why do you think that happens?
- For GPOa and GPOb: What logistical or IT issues have come up when trying to integrate scanning into routine antenatal care? How have you worked around these barriers? Are these workarounds sustainable in the long term?

When devices are conveniently stored and charged, scanning feels more seamless and gets done more often.

- In a site where scanning is working well, how are logistics managed? Is there a 'best practice' model?

When CHWs or clinicians juggle multiple responsibilities, scanning gets deprioritised unless it is seamlessly integrated into workflows.

- For GPOa and GPOb: How do doctors balance scanning with other antenatal duties at this successful site?

Wrap-up question

- If you could change one thing to make scanning easier or more sustainable in your clinic, what would it be?

Supplementary Material 7. Observational fieldnote template

Phrasing guide when discussing programme:

We want to do better care for people in your community with rheumatic heart disease. We have a new way of doing an ultrasound picture of our heart for children and pregnant women. We use a handheld scanner about the same size as/looks a bit like a mobile phone. The ultrasound can find rheumatic heart disease early. Then we can start treatment for the best care. We want to work together with people here. We want to find the best ways that people think will make this heart ultrasound check up work in this place. When we find the best ways, we can share it with other communities and other health services.

Discussion points and observations guide

- Who are you having the discussion with? e.g. role in community/clinic
- What are their ideas for the programme?
- Why will it work? (probing here for mechanisms e.g. ‘trust’, ‘confidence’, ‘knowledge’)
- How will it work? (what are the ‘steps’ involved? e.g. school screening, when, consent)
- How will you know it works? (probing here for indicators e.g. attendance)
- What assumptions are being made? (e.g. people get their check-up, secondary prophylaxis is available)
- What do you hope the program will achieve? (outcomes)

Supplementary Material 8. NoMAD survey

#### Survey instructions

This survey is designed to help understand how to integrate NEARER SCAN into usual care. It has three parts. Part A asks a few questions about you and your role. Part B includes three general questions about NEARER SCAN. Part C contains statements about NEARER SCAN with options to agree or disagree; where a statement is not relevant to you, please indicate why. Please select the option that best fits your experience for each question. The survey takes approx. 5–10 minutes.

#### Part A: About yourself

| 1. How many years have you worked for this clinic? | < 1 yr | 1–2 yrs | 3–5 yrs | 6–10 yrs | 11–15 yrs | > 15 yrs |
|---|---|---|---|---|---|---|
| Please tick one option | ○ | ○ | ○ | ○ | ○ | ○ |

| 2. Your job category in relation to NEARER SCAN | Please select one |
|---|---|
| ○ | I take the images of the heart using the handheld device (scanner). |
| ○ | I review the images off-site (reviewer). |
| ○ | I am a clinic manager at a site where NEARER SCAN is being implemented. |
| ○ | I am a GP/nurse/community health worker/other involved in RHD care but not scanning or reviewing for NEARER SCAN. |

#### Part B: General questions about the programme

##### How used to NEARER SCAN do you feel?

Still feels very new — Feels completely normal

0 1 2 3 4 5 6 7 8 9 10

##### Do you remember that you can do (if scanner), or check in about (if manager) NEARER SCAN everyday?

Not at all — Somewhat — Completely

0 1 2 3 4 5 6 7 8 9 10

##### Do you think you will be able to remember you can do NEARER SCAN everyday?

Not at all — Somewhat — Completely

0 1 2 3 4 5 6 7 8 9 10

#### Part C: Detailed questions about the programme

| Statement | Strongly agree | Agree | Neither | Disagree | Strongly dis-agree | Not relevant to my role | Not relevant at this stage | Not relevant to program |
|---|---|---|---|---|---|---|---|---|
| I can see how NEARER SCAN is different from other work. | ○ | ○ | ○ | ○ | ○ | ○ | ○ | ○ |
| Staff in this clinic have a shared understanding of the purpose of NEARER SCAN. | ○ | ○ | ○ | ○ | ○ | ○ | ○ | ○ |
| I understand how NEARER SCAN changes the way I do my work. | ○ | ○ | ○ | ○ | ○ | ○ | ○ | ○ |
| I can see the value of NEARER SCAN for my work. | ○ | ○ | ○ | ○ | ○ | ○ | ○ | ○ |
| There are key people who help lead NEARER SCAN at the clinic and get others involved. | ○ | ○ | ○ | ○ | ○ | ○ | ○ | ○ |
| I see taking part in NEARER SCAN as part of my role. | ○ | ○ | ○ | ○ | ○ | ○ | ○ | ○ |

| Statement | Strongly agree | Agree | Neither | Disagree | Strongly disagree | Not relevant to my role | Not relevant at this stage | Not relevant to the program |
|---|---|---|---|---|---|---|---|---|
| I'm happy to try new ways of working with others to use NEARER SCAN. | ○ | ○ | ○ | ○ | ○ | ○ | ○ | ○ |
| I will continue to support NEARER SCAN. | ○ | ○ | ○ | ○ | ○ | ○ | ○ | ○ |
| It's easy for me to fit NEARER SCAN into my work day. | ○ | ○ | ○ | ○ | ○ | ○ | ○ | ○ |
| NEARER SCAN makes it harder for people to work well together. | ○ | ○ | ○ | ○ | ○ | ○ | ○ | ○ |
| I have confidence in other scanners to use NEARER SCAN well. | ○ | ○ | ○ | ○ | ○ | ○ | ○ | ○ |
| When a scan needs to be done, others expect it is me that does the scan. | ○ | ○ | ○ | ○ | ○ | ○ | ○ | ○ |
| We get enough training to help us use NEARER SCAN. | ○ | ○ | ○ | ○ | ○ | ○ | ○ | ○ |
| We have all the equipment we need to do NEARER SCAN well. | ○ | ○ | ○ | ○ | ○ | ○ | ○ | ○ |

| Statement | Strongly agree | Agree | Neither | Disagree | Strongly dis-agree | Not relevant to my role | Not relevant at this stage | Not relevant to the program |
|---|---|---|---|---|---|---|---|---|
| Management supports NEARER SCAN. | ◯ | ◯ | ◯ | ◯ | ◯ | ◯ | ◯ | ◯ |
| I know there are updates or stories about how NEARER SCAN is going. | ◯ | ◯ | ◯ | ◯ | ◯ | ◯ | ◯ | ◯ |
| Most people at work agree NEARER SCAN is worth doing. | ◯ | ◯ | ◯ | ◯ | ◯ | ◯ | ◯ | ◯ |
| I value the effects that NEARER SCAN has had on my work. | ◯ | ◯ | ◯ | ◯ | ◯ | ◯ | ◯ | ◯ |
| Feedback about NEARER SCAN can be used to improve it in the future. | ◯ | ◯ | ◯ | ◯ | ◯ | ◯ | ◯ | ◯ |
| I can change how I work with NEARER SCAN. | ◯ | ◯ | ◯ | ◯ | ◯ | ◯ | ◯ | ◯ |

Supplementary Material 9. Costing workbook

The full costing workbook has been published open access for transparency and is available at:

https://doi.org/10.5281/zenodo.17611625.

Supplementary Table 1. Training and supervision dose by site

**Table** Training and supervision dose by site

| Month | Site 1 | Site 2 | Site 3 | Site 4 | Site 5 |
|---|---|---|---|---|---|
| 1 | | | | Training | Training |
| 2 | | Training | Training | | |
| 3 | | | Visit | | |
| 4 | Training | | | | |
| 5 | | | | Visit | |
| 6 | | | | | |
| 7 | | Visit | | | |
| 8 | | Event(off-site)[1] | Event | Event(off-site) | |
| 9 | | | | Training[2] | |
| 10 | | | Visit | Visit | |
| 11 | Visit | | Visit | | |
| 12 | | | | | Visit |
| 13 | | Event | Visit | Visit | Visit |
| 14 | | | Event | | Visit |
| 15 | | | | | |
| 16 | | | | Visit | |
| 17 | | | | | |
| 18 | Visit | | | Visit | |
| 19 | | | | | |
| 20 | | | | | |
| 21 | Workshop[3] | Workshop | Workshop & Event | Workshop | Workshop |
| 22 | | | | | |
| 23 | | Visit | | | |
| 24 | | | | | |

[1]Some scanners at site joined regional mass screening week with expert cardiologists and sonographers.
[2]Staff at additional facility in Site 4 trained.
[3]Community-of-practice workshop where scanners gathered from all five sites at a central location.

Supplementary Table 2. Finals CMOCs

| Final ID | Draft IDs | CMOC | Phase 1 | Phase 2 | Phase 3 | Substantive theories |
|---|---|---|---|---|---|---|
| 1 | P2.24 | When multiple staff are trained as scanners and visibly scanning (C), it may be considered normal work within the clinic (O), because seeing multiple colleagues scan makes it a shared expectation. | NA | "Multiple people to solidify that this is what the health practice does, [create] more conversations about it." (FGGP4)<br><br>[When asked directly if people see scanning as a clinic activity or an individual project] "Yeah, basically [the other clinic staff] just leave it up to us with the scanning, to figure it out and all that." (FNCHW1) | Site 3, which was the only site with multiple active scanners (>=3 staff completing >=5 scans across >=3 months), also had the highest mean response for the NoMAD item measuring shared understanding among clinic staff (4.44), compared with 3.0–4.2 at other sites. This supports the idea that visible participation by multiple staff reinforces a shared understanding of scanning across the clinic. | Normalisation process theory[62], complex contagions theory[64] |
| 2 | P2.41 | When staff with authority (e.g. GP or senior nurse), time capacity, and a genuine interest in scanning, are trained as scanners (C), they become personally invested in the programme (M), and advocate for changes to clinic processes that support scanning (O). | NA | "I emailed [the midwifery manager] to say 'I'm doing scanning can we make it part of the normal process?' And they said 'yeah, sounds like a good idea', and then with the midwives it kind of kicked off from that. So it was contacting managers." (GP5)<br><br>"Nurse 3 was trying to do the [scanning] roster [for antenates]... tried to call a meeting." (Nurse3(2))<br><br>"At site 3, forwarded text from Nurse 3 to team member: Nurse 3(3) was [rostered yesterday]... I will chat to them tomorrow. I've been sitting in for two weeks and there ain't no way I could have done it." (OFN3) | NA | NA |

| Final ID | Draft IDs | CMOC | Phase 1 | Phase 2 | Phase 3 | Substantive theories |
|---|---|---|---|---|---|---|
| 3 | IPT5, P1.6, P2.23 | When FNCHW scanners have a nurse or GP scanner buddy to scan with (or call on if needed) (C), they are more likely to scan (O), because they feel supported (M). | "[FNCHW3] loves 'sticky stamping' with [Nurse3], so when [Nurse3] is here, [FNCHW3] has far more confidence so it's an example of having confidence when you've got the right buddy with you." (CM3)<br><br>"I've got [GP5] to relate to... we've got two sets of eyes... we can both learn off one another... it's useful when you get stuck." (FNCHW5)<br><br>"When the doctor is not here and it's just me and [FNCHW2(2)], we kind of get lost in doing it really properly with the science so we do it when the doctor is here to guide us." (FNCHW2) | know [FNCHW3] and I can go out in pairs, partner and go out [to scan]." (Nurse3)<br><br>"It's nice just giving little bits of feedback to each other." (GP5)<br><br>"But it's just me, so I don't... you don't really have anyone to bounce off." (FGNurse5)<br><br>"[FNCHW5] is really good on the scanner but they often get me, we do our scans together." (FGGP5) | From qualitative data there were four FNCHWs with GP/nurse buddy pairs. They completed a median of 20.5 scans (IQR: 18.8–23.5), compared to a median of 9 scans (IQR: 3.0–12.5) among those FNCHWs without. | NA |

| Final ID | Draft IDs | CMOC | Phase 1 | Phase 2 | Phase 3 | Substantive theories |
|---|---|---|---|---|---|---|
| 4 | P1.13, P2.29 | When cardiac sonographers visit a site for ongoing support (C), scanners are more likely to scan (O), because expert presence makes them feel confident (M1) and that scanning is acceptable to prioritise (M2). | "Good thing is I'm hoping that come [the cardiac sonographer's] visit, the hands on, and them being around [will help]." (CM2) | "Definitely helped with [cardiac sonographer] coming out to [Site 4] twice now, just for two days or so and lifts up confidence and gets the momentum going again." (FGGP4)<br><br>"It was really good when [cardiac sonographer] came back... cause in this you need to lean on something, like that, they keep us straight." (FNCHW2)<br><br>Talking about scanning [with] cardiac sonographer today: FNCHW immediately said it is much easier, why? 'Because she can help me and I can ask her things.' (OFN3)<br><br>"[Other scanner at Site 5] is confident when [cardiac sonographer] visits and then their confidence drops." (GP5) | Scanners were significantly more likely to perform at least one scan on days when an expert scanner was present compared to days without an expert ($OR = 18.15$, 95% CI: 10.87–30.26, $p < 0.001$).<br><br>The average number of scans on expert visit days was also approximately 50 times higher than on non-visit days (3.58 vs. 0.07 scans per day). | COM-B theory[144], self-efficacy theory[138] |
| 5 | P1.45 | When scanners have had few opportunities to practice (C), they are less likely to scan without a visiting sonographer present (O), because skill decay over time reduces their confidence (M). | "I still got more to go... we have to do it more than once a week to really get it inside our head." (FNCHW3)<br><br>"I did a few scans on my own, but I think I'm lost at doing it a bit." (FNCHW2) | "I was going to say practitioner confidence, I think that's a big thing. FNCHW4 is very capable at scanning but hasn't done very many recently, and therefore she's probably lost her confidence. Would definitely ask me instead, if I suggest to her, she'll go 'oh no no no you do it' because she's kind of forgotten and she's lost confidence and it's that getting that momentum and that confidence and building that up. It's not so much the patient's confidence that is the issue it's the practitioners." (FGGP4)<br><br>"[From a cardiac sonographer about a scanner] they were very rusty and needed lots of instruction." (OFN) | As above | Ebbinghaus learning curve[137] |

| Final ID | Draft IDs | CMOC | Phase 1 | Phase 2 | Phase 3 | Substantive theories |
|---|---|---|---|---|---|---|
| 6 | P1.40, P2.81, P2.82 | When supervision for FNCHW scanners is delivered off-site and remote supervision via any mode of communication is not a usual practice (C), engagement is limited (O), because FNCHWs see it as optional and deprioritise it in favour of more immediate tasks (M). | "We just haven't managed to get any good conversation [about scanning] on WhatsApp yet." (OFN)<br><br>"In the WhatsApp group, like we all get the same message, but then I'll be the only person that responds." (Nurse4)<br><br>"In today's meeting we were talking about how the WhatsApp group hasn't really been engaged with by people here." (OFN) | "[Cardiac sonographer] We need to keep encouraging communication from the site end... we have really tried to make the 'check-ins' work over the last year – emails, SMS, calls, and Zoom." (OFN)<br><br>"[Cardiac sonographer B] I am more than happy to provide feedback on image quality, check if scans are on Tricefy and troubleshoot downloading if someone reaches out to me. But as [cardiac sonographer A] said, checking in with the non-experts... has not made any apparent difference in the past." (OFN)<br><br>"[Other implementation team member] I am surprised if checking in with [scanners] makes no difference, this is different to my experience." (OFN)<br><br>"And do you think that [the reason the FNCHWs aren't using WhatsApp for support as much is] because it maybe feels a little bit less familiar? You're not as used to using WhatsApp as the doctors might be?" (Interviewer) "Yeah, I'm not a WhatsApp fan. I don't know about the others, I don't know why they're not engaging." (FNCHW2)<br><br>"[From expert scanner who has had success with remote support]: Chat with [scanner] tonight, on WhatsApp, she has an idea for how to get scanning happening in [her site]." (OFN) | NA | Street-level bureaucracy[63] |

| Final ID | Draft IDs | CMOC | Phase 1 | Phase 2 | Phase 3 | Substantive theories |
|---|---|---|---|---|---|---|
| 7 | P1.41, P2.84 | When scanners seek remote support and receive prompt advice (C), it reinforces the clinical value of the programme (M), which increases their willingness to scan (O). | NA | "Having the ability to have [support] on the WhatsApp group and being able to get that feedback when you're doing it, knowing that it's there and the response times are very rapid so we get feedback really quickly. That's all very, very helpful." (Nurse3)<br><br>"Makes us feel that there's someone out there that we can just bounce it off." (Nurse3(2))<br><br>"I find the text feedback like the WhatsApp group just such a support. To be able to get the cardiologist's opinion on something like you know that day like just gives you so much more confidence in how useful [scanning is] so you want to do it more." (GP5)<br><br>"I can text through to the name and [the cardiologist] is awesome, you can get an answer within minutes of what they think of the scan. I think just having that support and being well organised I guess makes it feel like you are not just doing these scans for no reason." (FGGP5)<br><br>"There's been good communication through the WhatsApp." (FGGP4) | NA | NA |

| Final ID | Draft IDs | CMOC | Phase 1 | Phase 2 | Phase 3 | Substantive theories |
|---|---|---|---|---|---|---|
| 8 | P1.24, P2.55 | When FNCHW and nurse scanners have dedicated time set aside for scanning (e.g. screening events) (C), more scans are done (O), because this protected time within their workflow allows them to prioritise it (M). | "It is about carving out dedicated time [for scanning]." (CM3)<br><br>"Even if we could spend an hour doing RHD scanning during the week. We finish off but if we could spend an hour at five that'll give us time to do some scanning, if they could come up with that routine." (FNCHW3)<br><br>"Set times so you can scan and you can just sit in here and scan everyone that comes in." (FNCHW3(2))<br><br>"I think definitely having screening days is a good idea." (CM5) | "Even if we had a day a week, or even an afternoon a week, but I just know that if we didn't have any additional capacity, I would be in the back of my mind thinking, shit, I've got to pick up this kid, oh my gosh, I've got to do this. Oh god, I cannot forget this." (Nurse3)<br><br>"Allocate a time, a day, that is strictly a blockout day." (FGNEGP4)<br><br>"The scheduled events are much easier for us. It also frees up a bit of time for us because there's often some allocation of clinic staff." (Nurse3(2))<br><br>"It's purely a capacity issue for opportunistic, it's not lack of will, it's purely capacity." (Nurse3(2)) | At Site 3, the only site to hold regular events with scanning, on event days scanning averaged 8.6 scans per day compared to 0.1 scans per day on non-event days. | NA |
| 9 | P1.33, P2.53 | When FNCHWs can scan children who are already gathered (e.g. at schools or youth programmes) (C), more scans are done (O), because with the logistical responsibility removed, scanning feels like a manageable task (M). | "All the kids they're all going to be in one spot. That's fine like having that event or something for us to work with to go and do it is a lot easier. We just don't have the resources to organise something ourselves." (CM2)<br><br>"I'm just hoping maybe after the break, we can if we can just go to school and do it there with them." (FNCHW2) | "Yeah, we see at the last [public health scanning event], when we were at the Youth Center, they put it on Facebook, and then some of the girls that walked in 'oh, we seen it on Facebook. Yeah, that's cool.' " (Nurse3)<br><br>"We had a station for heart scans and we went up during [a local youth] programme, like with the kids sports, and we were just getting like two or three [scans] at a time." (FNCHW2)<br><br>"I think doing those kind of focused scanning days would be good. Like if I could even go up to the school next semester, yeah, we can see if we can organise like a day at the school where everyone gets scanned." (Nurse5) | As above | Invisible work theory[164] |

| Final ID | Draft IDs | CMOC | Phase 1 | Phase 2 | Phase 3 | Substantive theories |
|---|---|---|---|---|---|---|
| 10 | P1.30, P2.51 | When a clinic has a consistent manager and time to build a working relationship with the external programme team (C), the relationship fosters a sense of shared commitment (M1) and accountability (M2), which drives the manager to make practical changes to support scanning (O). | "Having the Menzies partnership as a long-term thing, [chief investigator A] and [chief investigator B] being the paediatricians here, and [one of the other investigators] as a long-termer as well. So there's this constant and continuous flow, flow of people who have a real strong interest in this space [which helps]."<br><br>"I said to FNCHW, we need to push [NEARER SCAN], you have to do it, so that first week they go in to have a go with it... but we need to start doing this." (CM2)<br><br>"I got in touch with [external youth organisation] last week and was like 'buddy, we need to get some kids for [FNCHW2] and [FNCHW2(1)] to get some numbers, is there any way we can combine it with the school holiday?" (CM2) | "Knowing that you guys are coming out [to visit the site] is also like crap, let's get sorted. Like they are coming out we need to get this done." (CM2)<br><br>"It's fully supported. Oh yeah, we just lead the way." (FGNurse3) | Sites with consistent clinic leadership (Sites 2 and 3) reported the highest scanner-perceived managerial support in the NoMAD survey (4.3 and 4.2). In contrast, support was lower at sites with leadership changes, such as Site 4 (2.3) and Site 5 (3.0). Site 1 showed an initial high level of support, which declined with manager turnover (3.5 by late implementation). | Normalisation process theory[62] |

| Final ID | Draft IDs | CMOC | Phase 1 | Phase 2 | Phase 3 | Substantive theories |
|---|---|---|---|---|---|---|
| 11 | P2.115 | When clinics experience high manager turnover and new managers face competing demands without clear guidance or working relationships with the external programme team (C), the resulting ambiguity (M) means manager support for scanning tends to remain nominal (O). | 'I missed out who got trained, as well." (CM4)<br><br>I think he had a bit of an issue with uploading the echos, I'm not sure how that's going." (CM5) | I couldn't even tell you honestly if they are doing any echos, you know, echos or anything at the moment as part of the NEARER SCAN programme, I don't, I'm not even sure that they are." (CM1)<br><br>To be honest, I didn't get any handover [about the programme]... maybe the new programmes run by external stakeholders are not identified as a priority, I think that could be a reason why it's not mentioned." (CM4)<br><br>Yes they [the clinic manager] keep changing they don't say why." (FNCHW4(2))<br><br>[Having new managers has been disruptive] because they don't even know what the NEARER SCAN is." (FNCHW1)<br><br>Manager has been on sick leave and then we had an interim manager and then another interim manager. So I think it's one of the big issues." (GP5)<br><br>I know that [previous manager] is [supportive]. You know, really on for this but [I'm] not quite sure about the other managers, yeah, whether or not they allow us that time. I know [the previous manager] would but she's not in that role anymore." (FNCHW1(2))<br><br>It's just, you know, crisis management day-to-day rather than having a bigger picture understanding of how things can work well." (GP4, reflecting on acting as CM4) | NA | NA |

| Final ID | Draft IDs | CMOC | Phase 1 | Phase 2 | Phase 3 | Substantive theories |
|---|---|---|---|---|---|---|
| 12 | P1.8, P2.38 | When clinic managers mention the programme in staff team communications (e.g. morning team meetings, celebrating scanning efforts) (C), FNCHW scanners are more likely to prioritise scanning (O), because it signals that scanning is a legitimate clinic activity (M). | "We spend a lot of time advocating for these things in morning meetings and just day to day." (CM3)<br><br>"Afternoon tea with the others in the clinic, clinic manager, AHPs, nurses to celebrate FNCHW2 [after finishing training]. All very proud and supportive, FNCHW2 saying 'If you guys see someone for an appointment let me know to do a scan."' (OFN)<br><br>"[At site 4] asked for show of hands who was aware of NEARER SCAN. Out of 13 staff, 5 aware – all [local Aboriginal people]. Contrast with [Site 3] meeting where manager knows all scanners." (OFN4) | "FNCHW3 introduced themselves [as a scanner] at all staff morning meeting after graduating and receiving a scanner shirt." (OFN)<br><br>"[When asked what we could do to help with scanning implementation at site 4] 'Normalising the idea of scanning within the practice so that the GPs, and the nurses, and the CHWs, all know that we are doing this and this is why we are doing it and it becomes a normal part of what we do."' (FGGP4) | In the NoMAD results, managerial support had a significant, weak positive correlation with shared understanding: ($\rho$ = 0.37, p = 0.032). | Role legitimacy[162]; social norms theory[246]; normalisation process theory[62] |

| Final ID | Draft IDs | CMOC | Phase 1 | Phase 2 | Phase 3 | Substantive theories |
|---|---|---|---|---|---|---|
| 13 | P1.17, P2.45 | When clinic managers emphasise the benefits of scanning for the community (C), it resonates with FNCHWs' commitment to caring for the community (M), which makes them more willing to participate in scanning (O). | "[Text message from CM2]: I'm so so proud of you [FNCHW]. I hope you're proud of yourself... It's so awesome to see these guys do something that can have such an impact on the people of [Site 2] and their lives. Keep up the good work." (OFN)<br><br>"Thank you all for this opportunity and new skill we can now have in our local clinic for our community." [Message from FNCHW2] (OFN) | "My name is [FNCHW3], I'm a community health worker, and also I'm a NEARER SCANNER, and I'm so excited to do some scans today, wearing this new deadly shirt I've got." (OFN3)<br><br>"And it's something that they can achieve on their own. Something super important. For them, 'I did this'." (CM2)<br><br>"I feel proud to be [scanning], looking at that kid... looking at his face makes you feel emotional." (FGFNCHW2) | NA | NA |
| 14 | IPT2, P1.31, P2.52 | When clinic managers regularly check in with scanners about their scanning (C), it signals that scanning is important amid competing demands (M), which reinforces that scanning is legitimate to prioritise (O). | NA | "Guys do you reckon, you can smash out 15 [scans]?" (CM2)<br><br>"It's the next step, which is how does the manager actually build that into their programme and is that being done? Is there regular check-ins saying how you're going [to the scanners]? Have you got a couple of scans this week? Or whatever like that, just regular check-ins, I think that's the next step. It's not being done here." (FGGP5)<br><br>"Now there is an expectation from the clinic that all women will get a scan. Once it becomes an expectation of the clinic it happens." (FGGP6) | NA | Role legitimacy[162] |

| Final ID | Draft IDs | CMOC | Phase 1 | Phase 2 | Phase 3 | Substantive theories |
|---|---|---|---|---|---|---|
| 15 | P1.34, P2.69, P2.70, P2.71 | When the practical tasks needed to keep scanning operational (e.g. charging, locating the device) are both accessible to and expected of all scanners (C), it signals that scanning is a shared responsibility (M), making scanning activity more likely to be evenly distributed across trained staff (O). | "So [FNCHW5] normally has his in his room." (Nurse5)<br><br>"The scanner usually stays in here, Nurse5 is in the [building] out there. So if she wants to scan, she has to come grab the scanner." (GP5)<br><br>"[FNCHW2] suggests Lumify can be kept in [the clinic manager's] office, to access when using." (OFN2)<br><br>"[FNCHW2 and FNCHW(2)] share the one scanner. There are no issues with either of them accessing the scanner. They keep it in the manager's office because their consult rooms are used for on call which means there can be numbers of other people in the room. Manager's office is easily accessible." (SR2) | "We've had issues with the Lumify not being charged in the beginning, but they now live in our locked office and have their own little set of cable cords, and it seems to be working." (FGGP6)<br><br>"Now [the device] lives in our locked office... we have them all in the same office." (FGGP6(2))<br><br>"[IT support person] is amazing and has [the devices] plugged in in his office [which is accessible]." (OFN)<br><br>"We will always find one, one of us always finds one." (FGNENurse3)<br><br>"[Site 5 scanner] stated that after the initial training they felt confident with scanning but has lost that through not being able to obtain the Lumify [from other scanners]." (OFN5)<br><br>"It's in my room." (FGGP4)<br><br>"The one I am using at the moment was in Nurse5's room but she was not scanning so now it's in my room." (FGGP5) | Site 5, which had the most reports of certain scanners feeling they lacked access to scanners, recorded the highest Gini coefficient (0.48), indicating scanning was concentrated to one scanner. In contrast, Site 3, which had a strong culture of sharing devices, had the lowest Gini coefficient (0.10), suggesting scanning was more evenly distributed. | Invisible work theory[164] |

| Final ID | Draft IDs | CMOC | Phase 1 | Phase 2 | Phase 3 | Substantive theories |
|---|---|---|---|---|---|---|
| 16 | P1.35 | When devices aren't ready for immediate use because the operational tasks (e.g. charging) have not been done (C), scanners may abandon their intention to scan (O), because of the frustration that arises when the expectation of scanning as a ready-to-go task is undermined (M). | Device required charging during which time FNCHW4 continued her clinical work. I notified FNCHW4 when there was sufficient charge on the Lumify however she appeared to have lost interest in heading out to community. (SR4) | "These machines were being a bit dodgy with the charging... there's a few times we had some issues with them, they'd been on charge for days or hours and then they weren't fully charged." (Nurse3)<br><br>"Knowing that the equipment's working and charged and it's already to go, that speeds things up." (GP4)<br><br>"[Interviewer] When devices are conveniently stored and charged, scanning feels more seamless and gets done more often. [Sounds of agreement]." (FG of scanners)<br><br>"I don't use it yeah, because not on charge." (FGNurse3) | NA | Invisible work theory[164] |
| 17 | P1.1, P2.9 | When other clinic staff, especially GPs, refer patients to FNCHW or nurse scanners for a scan (C), it acts as a social nudge that legitimises (M) the prioritisation of scanning (O). | "[The GP] is pretty on to it, he pulls us up, asks us, or even brings patients to us." (FNCHW5)<br><br>"I remember the last time [the paediatrician] was here, he called me and he asked me if I had some spare time to go with him to do a heart scan on a patient. He's really good at doing that, very supportive. He likes to help us out to learn more about scanning." (FNCHW3) | "We're now getting GPs coming and saying, 'Hey, I've just got a kid can you just do a quick scan?"' (N3)<br><br>"Sometimes doctors will come 'can you do the scan?' Or, a nurse comes, 'can you do the scan?"' (FNCHW2)<br><br>"[Scanning is] not done, we don't get to do any scanning, [but] when [the paediatrician] was here, the senior people, send the text message 'do you or anyone wants to do scanning come down."' (N3(2)) | NA | Nudge theory[247]; role legitimacy[162] |

| Final ID | Draft IDs | CMOC | Phase 1 | Phase 2 | Phase 3 | Substantive theories |
|---|---|---|---|---|---|---|
| 18 | P2.13, P2.18 | When scanning is visible to other clinic staff (e.g. scanning events) and is explicitly supported by line managers (C), other clinic staff are more likely to refer patients for scans (O), because scanning is collectively seen as an expected clinic activity (M). | NA | "Nurse at site 3 said it [the reason they are now getting scan referrals when they hadn't previously] probably is because of the promotion during [screening event] and new clinic staff seeing it more as standard practice." (OFN)<br><br>"[Referrals] just came with time, [once the other staff] were aware that I like to scan all the kids, because they would call me in for whatever reason and I would then offer a scan, then they would be like 'oh okay this is something that the GP is keen on doing.' So I think over time they are like 'oh, the GP might want to scan this kid, he hasn't had a scan already."' (GP5)<br><br>"I emailed the midwifery manager to say 'I'm doing scanning can we make it part of the normal process?' And they said 'yeah, sounds like a good idea', and then the midwives kind of kicked off from that. So it was contacting managers." (GP5)<br><br>"Getting that support from higher up manager. I think yeah, it was definitely part of it. I think they were very keen to get on board anyway, but yeah, it was definitely part of it, the manager." (GP5)<br><br>"It's to do with the doctors that know we do exist, but we don't get much referrals from other staff." (Nurse3) | This context can be clinic-wide, as seen at Site 3 with the highest NoMAD scores across the sites in both shared understanding (4.44) and managerial support (4.44), or within specific teams in a clinic as demonstrated by the qualitative accounts of a GP and the midwifery team at Site 5.<br><br>The *and* of the context is reflected in how at Site 2 with the second highest scores of both shared understanding (4.20) and managerial support (4.25) but did not have regular visible scanning occurring like in Site 3 which had regular event scanning. | Normalisation process theory[62] |

| Final ID | Draft IDs | CMOC | Phase 1 | Phase 2 | Phase 3 | Substantive theories |
|---|---|---|---|---|---|---|
| 19 | P2.64 | When FNCHWs' workflows are dominated by delegated or KPI-linked tasks and there are no clear social or formal expectations to scan (C), it is hard to prioritise (O), because it feels optional (M). | "Mostly I'm going work for the nurse." (FNCHW3)<br><br>FNCHW at Site 3 performed 2 scans and was then needed in liaison role. (OFN)<br><br>"Busy doing other stuff as well with short staff and with driving." (FNCHW2)<br><br>"I haven't scanned anyone yet. Always busy, flat out, working, need to catch up on what we do." (FNCHW(2))<br><br>"Just finding that extra 10 minutes to do echoes is probably a bit daunting when you've got lots of competing priorities." (CM5) | "Well, we've got our own jobs... we're not recognised [for] that RHD scan that you said, okay, so it's in our time." (FNCHW1)<br><br>"We're just getting told what to do here, there, and everywhere, you're not really valued." (FNCHW1(2))<br><br>"Compared to Site 6 where 'The organisation is expecting us to do it, there is a KPI on screening in everyone."' (FGGP6)<br><br>"How do you decide what work you do in the day? Even the cleaners may ask for help, and we help them to move furniture or something." (OFN in discussion with FNCHW)<br><br>"If [scanners] can't see that scanning is important or a legitimate bit of a part of your role, then the problem with that is that you've got all these other jobs that are going to be prioritised." (FGNurse5)<br><br>"But I guess the scans aren't... part of KPI's and... it's not part of the managers priorities and what they're overseeing [Nurse5] do, I guess. I think that the biggest barrier as to why they don't get as many numbers." (GP5) | A decline in scanners' scoring of the NoMAD item "It's easy for me to fit NEARER SCAN into my work day" from Phase 1 (4.29) to Phase 2 (2.56) suggests that scanning became less embedded in daily workflows over time. This may indicate that, without the initial expectations present immediately after training, scanning was harder to prioritise alongside other tasks. | Role legitimacy[162]; street-level bureaucracy[63]; normalisation process theory[62] |

| Final ID | Draft IDs | CMOC | Phase 1 | Phase 2 | Phase 3 | Substantive theories |
|---|---|---|---|---|---|---|
| 20 | P2.66 | When scanners have authority within the clinic hierarchy (e.g. GPs) they are trusted to manage their own workflow (C), so they are more likely to be able to prioritise opportunistic scanning (O), because this autonomy gives them the confidence to decide when scanning is appropriate (M). | NA | "[Interviewer: Do you feel like those competing responsibilities has a different impact for doctors vs FNCHWs? Is it easier for your to justify a scan?] "Yes probably, I am not expected to pick up patients from the waiting room, maybe they do feel more of a pressure if waiting room is full to get patients through, whereas in my role I can take my time." (FGGP6)<br><br>"[For FNCHWs, the manager looks at how many patients they are seeing. . . [whereas] it's me who decides how often I see which patients, so my workflow is in my hands." (FGGP6)<br><br>"A doctor can make it bit more independent decisions about what they do or what they need to focus on. . . [whereas] [Nurse5] has people kinda like watching those numbers [of KPIs]." (GP5) | NA | Professional authority[168]; street-level bureaucracy[63] |

| Final ID | Draft IDs | CMOC | Phase 1 | Phase 2 | Phase 3 | Substantive theories |
|---|---|---|---|---|---|---|
| 21 | P2.34, P2.35 | When there is a recall system for scanning that is visible to all staff (C), opportunistic scanning becomes easier to prioritise (O), because the recall provides visual prompts for action (M1) and makes scanning feel legitimate (M2). | NA | "We actually have a list of all our pregnant patients and we've incorporated the screening onto that. It's a column on an Excel spreadsheet. When someone comes in, we look and go 'oh, she hasn't had an echo, we'll do it!'" (FGGP6)<br><br>"We have a NEARER SCAN recall, 'Has had a scan', 'Is it reported?' it highlights red. That has worked with our antenatal patients because the midwives have put that recall on and ticked those boxes. So that's our spreadsheet." (FGGP5)<br><br>"Recall, something that pops up on the screen, makes it feel legitimate." (FGGP6)<br><br>"But to have that full [recall] list and then print it, these guys [FNCHWs at Site 2] are visual. And once you've knocked it out [saying] 'you're down 6 pages now, guys, look at us go, do you reckon you can knock off another page?'" (CM2)<br><br>"Recalls are a good idea, because people work off recalls." (FGGP6) | NA | Nudge theory[247]; role legitimacy[162] |
| 22 | P1.26, P2.58, P2.59 | When children present unwell for reasons other than suspected ARF/RHD (C), FNCHW and nurse scanners often choose not to offer scanning (O), because offering an optional check feels inappropriate when there are acute care needs (M). | "Most of the time they are sick already so people kind of just want to come in and get their treatment and go which is understandable. And so then it's quite hard to kind of like say 'oh well actually stay and do this as well', especially if it does take us a bit longer because I feel like I'm still not very comfortable at it." (Nurse4) | "I've been told we need to do opportunistic scanning for patients... it doesn't make sense to me because they're unwell... they've got something else that's more important to them at that time and then going and asking them if I can scan the child or the other children that are there, it's massive you know. It feels... [inappropriate someone else says] yeah it feels that." (FGNurse1)<br><br>"Yeah, no, if it's not going down that pathway [ARF workup], yeah, I wouldn't think about doing it." (FGNurse3)<br><br>"I asked her [other FNCHW at clinic 2] about opportunistic scanning. She didn't think she would do that unless a child presented with symptoms." (OFN) | NA | Street-level bureaucracy[63] |

| Final ID | Draft IDs | CMOC | Phase 1 | Phase 2 | Phase 3 | Substantive theories |
|---|---|---|---|---|---|---|
| 23 | P2.57 | When scanning is offered during a quick visit for a specific reason (e.g. bicillin) (C), participants seem less likely to agree (O), because they want to limit time in the clinic (M). | "They just want to come in do that [one thing], you know, their bicillin or whatever, they come in for that." (Nurse3)<br><br>"Some of them [say when declining scans] 'oh, we already, seen the doctor.' " (Nurse3) | "And sometimes [patients] come and they want to get their LAB and we are hopeful to offer a scan and they say I'll come back tomorrow, maybe later, maybe another day. So that's something it's not helpful." (FNCHW3)<br><br>"It's got to do with the patient willing to to have it done because... we do ask some [say] 'tomorrow' and we know that's a no." (Nurse3) | NA | Street-level bureaucracy[63] |
| 24 | P2.60 | When children become restless during a long appointment (C), scanners are less likely to offer a scan (O), because they want to finish core clinical tasks while the child is still engaged (M). | "They might engage a bit easier if it's just that they come in it takes 10 minutes, they just do the scan and that's done rather than in an appointment, which might already take an hour... I think what we're doing at the moment, well not I think, it's not really working, we are not getting them done." (Nurse4)<br><br>"Health checks are sometimes a luxury." (CM3)<br><br>"I find that school age check is already really long, both to the clinic, but also for the child and the family... the kids get bored and want to leave and the parents kind of get bored and want to leave, already like the volume of like how long but then adding [scanning is] one of the things that makes it quite hard." (Nurse4) | "When you've got really high flow rate of patients in the clinic in child health... you've got this incredibly narrow window to capture as much as possible before the child starts getting upset. The mother then says, 'that's it - done' and they go home... and then, of course, you're just looking down your priority list, and it's [scanning] just not there, right?" (Nurse3(2)) | NA | Street-level bureaucracy[63]; invisible work theory[164] |

| Final ID | Draft IDs | CMOC | Phase 1 | Phase 2 | Phase 3 | Substantive theories |
|---|---|---|---|---|---|---|
| 25 | P2.94 | When FNCHWs work in their own community and encounter eligible scanning participants where a cultural protocol prohibits interaction (C), they choose not to scan them (O), because they feel responsible for upholding cultural norms to maintain respect (M). | As above | "It does [stop us from scanning sometimes], it does yeah, cultural barrier." (FGFNCHW3)<br><br>"That why it's good to have me and FNCHW2(2), he does the male, I do the female. The cultural barrier is all about respect." (FGFNCHW2)<br><br>"Actually one barrier, I think we haven't actually said amongst us is we're all female scanners." (FGFNCHW1)<br><br>"Sometimes I feel uncomfortable scanning males...[and] poison cousins stops us from scanning yep, if it's a boy or boy and girl, yeah, we can't talk to each other or go near each other or touch each other because they're poisoned cousins." (FNCHW3) | NA | NA |
| 26 | P2.68 | When FNCHW scanners work in their own community and carry substantial emotional and family/community responsibilities (C), cumulative fatigue reduces their mental and physical bandwidth (M), which may lead them to deprioritise non-urgent tasks like scanning (O). | As above | "But sometimes... you go to work, [and it] make you feel extra tired, because sometimes work don't stop, you've got to still worry about heart problem and stuff like that [outside of work]. Humbug [a local term that means unreasonable or excessive demands from family or community] after work." (FNCHW3)<br><br>"I had to stop people from humbugging [FNCHW]... [the patients say to her] 'hey, I just want this. I just want my tablets. I just want this. I just wanna go.' It's non-stop... the constant humbug in here, and you're seeing 10 different people, and once that noise is gone, you don't feel as much pressure." (CM2)<br><br>"It's tricky for [FNCHW4]. She's been called in lots of different directions... and then has a lot of family, humbug, family responsibilities on top of that. So I think for her to even have some breathing space, never mind think about doing scanning. It's just not feasible." (GP4) | NA | Invisible work theory[164] |

| Final ID | Draft IDs | CMOC | Phase 1 | Phase 2 | Phase 3 | Substantive theories |
|---|---|---|---|---|---|---|
| 27 | P2.1, P2.2 | When GP scanners offer opportunistic scans in a busy clinic environment (C), the consent process may be quicker and involve less patient education (O), because GPs consider brief information sufficient for consent at the screening stage (M1), and patients may be more willing to consent (M2). | NA | "My consent and education is fairly brief. I will often just like come into the room and just ask do you mind if we just do a heart checkup?" (GP5)<br><br>"I feel when I am doing the scan I don't have time to explain what I am doing, what it is." (GP4)<br><br>"It is just a quick thing… I don't think it needs a lot of consent and explanation… if most of the times [the result] is going to be normal." (GP5)<br><br>"[In response to a GP from another site asking if they take the time to explain RHD at Site 6] A little bit, depends on the woman and how many people in the waiting room. Depends on how accepting, or health literate I guess, or high risk… so pretty variable." (FGGP6)<br><br>"I feel the same, it's about how it's sold and how it is talked about." (FGGP6(2))<br><br>"If AHPs are doing a lot of education that's not a bad thing because you know it's a good public health message and they can talk about it and their family groups. [But] if you're looking at building [scanning] as easy everyday thing, I think you can keep the consent and the education basic until actually there is [an abnormal finding]." (FGGP5)<br><br>"We were given resources and a video for the consent process, right? We found that there's no time for that and pregnant women don't seem to sort of want to seek out that basically, they accept the scan for their heart." (FGGP6) | 83% of GP scans occurred when no study team member was on-site (as a proxy for opportunistic scanning) vs 13.8% for FNCHWs. GPs were significantly more likely to scan during these times than FNCHWs (OR 29.7, 95% CI: 13.8–68.4, $p < 0.001$, Fisher's exact test). | Self-efficacy[138]; street-level bureaucracy[63]; professional authority[168] |

| Final ID | Draft IDs | CMOC | Phase 1 | Phase 2 | Phase 3 | Substantive theories |
|---|---|---|---|---|---|---|
| 28 | P2.4, P2.5 | When FNCHW scanners offer opportunistic scans in a busy clinic environment (C), the consent process may take longer and involve more patient education (O), because FNCHWs see it as their role to explain RHD and scanning in a relatable way (M1), and patients may feel more comfortable asking questions in the absence of language and cultural barriers (M2). | NA | "When they see us doing [scans] they get the idea of what's happening with our heart... some of them in the community doesn't know what's happening to them. They don't know how long they are having [penicillin injections] for. It's up to us to give them the feedback." (FGFNCHW1)<br><br>"We are not going making decisions and telling them that we're going to [scan]." (FNCHW3)<br><br>"It's good for them to see their own people doing stuff like this, they feel comfortable... you can giggle, you can laugh, you can talk about the scan, you show them their heart." (FGFNCHW2)<br><br>"[Doctors] don't really explain it properly. So most of the time, they'll ask for us to go sit in with them, so then we can explain it thoroughly instead of using scientific words." (FGFNCHW1(2)) | NA | NA |
| 29 | P2.7 | When FNCHW or nurse scanners have the opportunity to scan in schools but feel unsure about the consent process (C), they may choose not to scan (O), because they are concerned about doing the wrong thing (M). | NA | "But then what about their consent form [at the school]? Is that going to be a barrier? Last time there was a bit of a barrier." (FGFNCHW1)<br><br>"Nurse at site 5 needs consent forms to scan kids at school." (OFN)<br><br>"Consent is a big thing because you don't want to go wrong. It has to be an easier process... We try scan at the school but consent is an issue." (FGFNCHW1(2)) | NA | NA |
| 30 | P1.37, P2.73 | When image upload issues (e.g. internet connectivity or upload app errors) are frequent and unresolved (C), scanners may stop scanning (O), because they anticipate the effort won't deliver clinical value (M). | NA | "[Connectivity and upload issues have] been a big disincentive because it's been well I can do the scan, but I'm gonna have to take time to try and troubleshoot to get uploaded." (GP4)<br><br>"When we first started we had a lot of issues uploading, and that was definitely a barrier and that's now gone away, which I find helps me a lot, I'm not having to spend half an hour going into settings and testing it out." (FGGP6) | NA | Sociomateriality theory; invisible work theory |

| Final ID | Draft IDs | CMOC | Phase 1 | Phase 2 | Phase 3 | Substantive theories |
|---|---|---|---|---|---|---|
| 31 | P1.38, P2.74 | When image upload issues (e.g. internet connectivity or upload app errors) are frequent and unresolved (C), scanners may delay uploading (O), because repeated failures make delayed uploading an accepted norm, even if scanners know it's not best practice (M). | "I have done scans but I haven't uploaded any." (GP5) | "Sometimes scans have sat there for a long time and just not got uploaded because I haven't had time to talk to anybody... in the past it's been 'I can't be bothered to' it takes the time to do the scan, takes the time to upload. It's just gonna not be worth it? Can't face that." (GP4)<br><br>"Local scanners cannot be expected to spend the amount of time currently required to ensure images are being sent." (SR3) | NA | Street-level bureaucracy theory[63] |
| 32 | P2.76, P2.72 | When upload issues persist despite local attempts to troubleshoot (e.g. home WiFi or mobile hotspot), even when remote support is accessible (C), scanners usually wait for a sonographer visit (O), because remote troubleshooting feels like an added burden in an already busy workflow (M). | "Again, a lot of time was spent gaining wifi access and uploading studies." (SR1) | "[If upload issues] I'm going to have to message somebody and have to talk to somebody and it's just going to take time." (GP4)<br><br>"No scans sent from this site since Jan 24 (6 months) due to unable to access internet." (OFN2)<br><br>"The internet remains a huge barrier. A lot of time was spent [by me, cardiac sonographer] checking connections and checking uploading was happening." (SR3) | NA | Street-level bureaucracy theory[63] |

| Final ID | Draft IDs | CMOC | Phase 1 | Phase 2 | Phase 3 | Substantive theories |
|---|---|---|---|---|---|---|
| 33 | P1.42, P2.89 | When FNCHW and nurse scanners don't receive clear confirmation that scans have uploaded and will be reviewed by experts (C), they may start doubting whether the process will improve care (M), so they could stop scanning (O). | "I feel like my scans go into the abyss, how do I get results?" (Nurse4)<br><br>Quote from FNCHW – "Currently, without being able to upload the images it makes us frightened to scan because if we find any problem such as regurgitation, we can't go any further. We can only do half the process." (OFN)<br><br>"Trainees here said they are scared to take images because if it can't be uploaded (and thus reviewed) they feel bad taking the imaging in case it is a positive scan and it goes nowhere." (OFN) | "I was concerned that somebody wasn't looking at one that I had done with you. And so that caused me anxiety." (Nurse1)<br><br>"It's more about whether or not scanning is something useful and good for them to do, for their patients, who they are very motivated to help and care for... I think a big part of us getting this right is finding ways to make sure that when people get scanned, this results in a useful result that is good for the patient and the team looking after them. Having good internet will be a big game changer." (OFN)<br><br>"[Team member 1] It clearly makes the non-experts nervous if they aren't certain the scans will be seen and reported on. They worry they will miss something. [Team member 3]: And so stops them scanning." (OFN)<br><br>"Also I know one of the reasons [for low scan numbers] is because there are scans done from two months ago that we have been chasing results that we haven't got back to them. This is issue on our end that would definitely be impacting. I know I would stop scanning because what I tell people about it being looked at by a cardiologist and results in a few days isn't being followed through." (OFN) | NA | Street-level bureaucracy theory[63] |

Supplementary Material 10. Implementation toolkit

### Orientation to SPLASH scanning for RHD in communities

#### Is our clinic ready?

This brochure provides information for clinics interested in Rheumatic Heart Disease screening using a SPLASH scan. It draws on lessons from the NEARER SCAN study in five clinics across the NT and WA. The study highlighted what helps scanning to become part of routine practice and key factors that can support this process. Not everything needs to be in place from the start, as readiness can develop over time. Strong clinic leadership is important, and the Menzies team will provide support along the way.

#### Who to train as a SPLASH scanner

We encourage clinic managers to discuss the scan training plan with all staff to find the best fit. Consider who is interested, available, and likely to stay in the community—local health workers are often the most long-term. Training a long-term GP or senior nurse can provide additional support to scanners and some trainees found having a 'scanning buddy' helpful. Seek input from community elders, schools, and local groups about heart scanning as well. Look for local RHD Champions.

SPLASH:
Single Parasternal Long Axis acquisition with a Sweep of the Heart (the simplified scan used for RHD screening)

ASUM:
Australasian Society for Ultrasound in Medicine
(new online modules now created for SPLASH training)

##### Cultural sensitivity and awareness

- ☐ 1 Are there cultural considerations and sensitivities we should be aware of in relation to scanning in your community?
- ☐ 2 What is the clinic's approach to cultural safety in the workplace and in interactions with the community?

##### Clinic systems and scheduling

- ☐ 3 Could trainees have dedicated time to complete the ASUM online modules for SPLASH scanning? Is any additional support required (e.g. technical or language support)?
- ☐ 4 Could scanning time be scheduled periodically for training and afterwards? (e.g. screening events/ clinics)
- ☐ 5 Could a process be created where individual scanners or the health service is notified once scans have been reviewed so that scan results (both normal and abnormal) are acted upon?
- ☐ 6 Could SPLASH screening be included in the clinic KPIs or routine performance measures?

##### Knowing who and when to scan

- ☐ 7 Could a recall list be used to identify people who are due for a SPLASH scan?
- ☐ 8 Could the clinic work with local schools to develop a consent process for adding SPLASH scans to school screening?
- ☐ 9 Could all staff be informed about the process for referring people for a SPLASH scan?

##### Team support

- ☐ 10 Could scanning be discussed at existing regular staff meetings (e.g. morning huddles)?
- ☐ 11 Could the clinic schedule regular check-ins with scanners to talk about how scanning is going?
- ☐ 12 Could the clinic facilitate regular chats or visits from the cardiac sonographer to support the scanners?
- ☐ 13 Could the clinic manager develop a scanning handover plan in case of management changes?

##### Technical set up and accessibility

- ☐ 14 Does the clinic have reliable Wi-Fi or mobile internet for uploading scans? What support or infrastructure is needed to ensure scanners have consistent access?
- ☐ 15 Is there an accessible secure place to store and charge the devices?

menzies SCHOOL OF HEALTH RESEARCH

#### Maintaining scanner's skills

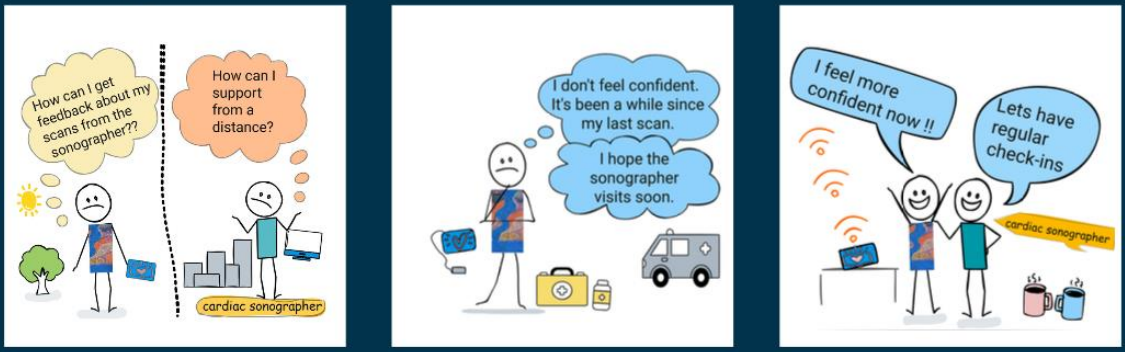


#### Timeline for training

| | 1 | 2 |
|---|---|---|
| **STAGE** | **Connecting to community and clinic** | **Set-up period** |
| **TIME** | Unspecified | 2–3 months |
| **FOCUS** | Relationship building | Planning & Toolkit |
| **KEY ACTIVITIES** | Schedule time for the Menzies team to meet and develop relationships and understanding of the community and its health service. | – Identify trainees<br>– Discuss readiness Toolkit & checklist<br>– Sonographer visit: introduce ASUM portal, scanning device, and training plan<br>– Trainees start ASUM online modules |

#### Clinic managers influence scanning

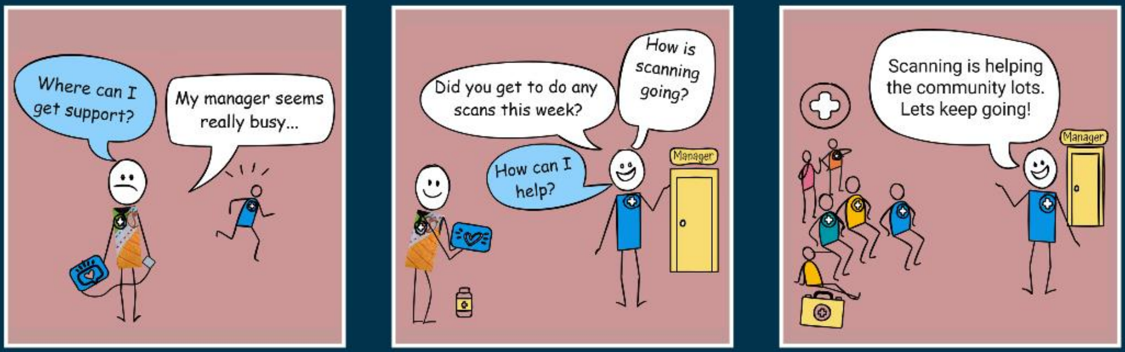

#### Screening events help with skills

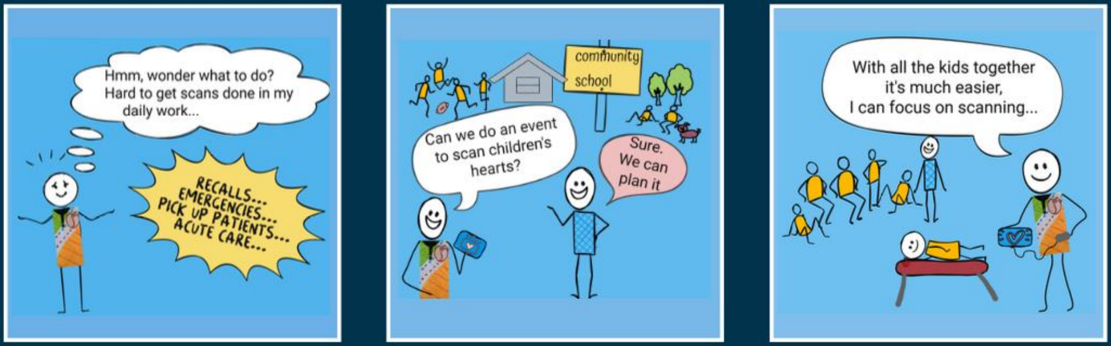


##### 3 On-site training

3–4 days

Hands-on learning

- In-person training with sonographer
- Review ASUM modules
- Practice scanning techniques
- Plan sonographer-supported screening events

##### 4 Local scanning

2–6 months

Independent scanning practice

- Begin day-to-day SPLASH scanning
- Upload scans for review
- Receive feedback
- Participate in sonographer-supported screening events

##### 5 Graduate SPLASH Scanner

Upon 100 scans

Certification & focus

- Graduate as a SPLASH scanner
- Receive ASUM qualification
- Plan ongoing scanning prioritising ages 5–20 yrs, pregnant women, ARF cases and priority 3 RHD follow-ups

#### Connecting scanning to workflows

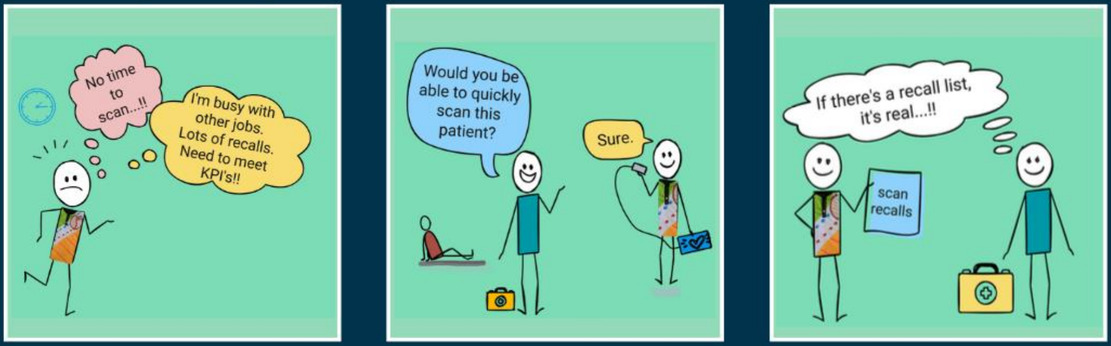

### Success Stories

*The following stories are real examples from the work we have done, showing how staff in different roles took practical steps to make scanning part of everyday work.*

At one clinic, an Aboriginal Health Practitioner created recalls in their clinic's medical records for yearly SPLASH scans for children aged 5–20 and currently pregnant women in their community. They helped run a scanning station during a Youth Sports programme, where they said they were able to do "two or three [scans in a row]." They also felt supported by visiting cardiac sonographers, noting that "they keep us straight." Importantly, local cultural systems were considered in helping who to choose to train, with male and female staff sharing responsibility: "That's why it's good to have me and [another scanner here], he does the male, I do the female....[it's] all about respect."

**Aboriginal and/or Torres Strait Islander health practitioner / health worker / community worker**

At one clinic, the manager actively supported the programme and motivated staff. They encouraged trained staff to scan, saying "we need to push [scanning]...we need to start doing this," and highlighted its benefit to the community: "it's so awesome to see these guys do something that can have such an impact on the people of [this community]." The manager also looked for opportunities to link scanning with community events, asking the organiser of a local school holiday program "is there any way we can combine [scanning] with the program?" Additionally, they coordinated visits from cardiac sonographers to provide hands-on support.

**Clinic manager**

**Nurse**

At a third site, a nurse helped organise a fortnightly SPLASH scan clinic for women in early pregnancy. They also organised scanning to be part of a larger community health promotion event. They noted that referrals for scans increased after this, likely because people saw scanning happening. The clinic staff began to see scanning as part of their everyday work. Overall, the nurse described the programme as "fully supported... we just led the way."

**General practitioner**

At another site, a GP became a strong champion for point-of-care SPLASH scanning. They often worked alongside an Aboriginal Health Practitioner to scan, saying it was good to give "little bits of feedback to each other." This GP helped make scanning everyday work in several ways. First, by setting up a referral process with the midwives ("I emailed [the midwifery manager] to ask, can we make it part of the normal process? And then the midwives "kicked it off"). Second, by encouraging colleagues to refer children for scans ("Over time they were like, 'oh, the GP might want to scan this child'"). And third, by sharing stories in morning meetings about how scanning improved care for patients, such as speeding up retrievals when severe valve disease was detected.

#### Deciding when to offer scans

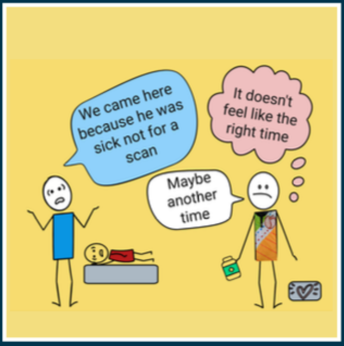


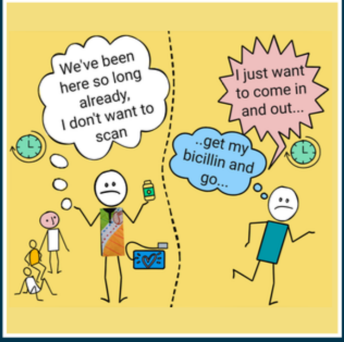


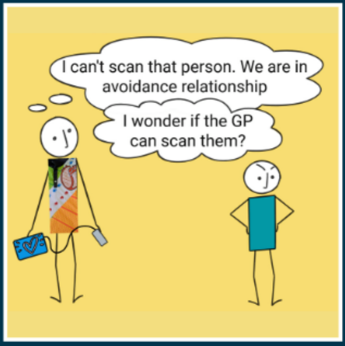


The contents of this brochure are solely the responsibility of Menzies School of Health Research and do not reflect the views of the Commonwealth

MRFF2041013
